# End-to-end prediction of clinical outcomes in head and neck squamous cell carcinoma with foundation model-based multiple instance learning

**DOI:** 10.1101/2025.01.22.25320517

**Authors:** Asier Rabasco Meneghetti, Marta Ligero Hernández, Jens-Peter Kuehn, Steffen Löck, Zunamys Itzel Carrero, Raquel Perez-Lopez, Keno Bressem, Titus K. Brinker, Alexander T. Pearson, Daniel Truhn, Sven Nebelung, Jakob Nikolas Kather

## Abstract

**Background:** Foundation models (FMs) show promise in medical AI by learning flexible features from large datasets, potentially surpassing handcrafted radiomics. Outcome prediction of head and neck squamous cell carcinomas (HNSCC) with FMs using routine imaging remains unexplored.

**Purpose:** To evaluate end-to-end FM-based multiple instance learning (MIL) for 2-year overall survival (OS), locoregional control (LRC), and freedom from distant metastasis (FFDM) prediction and risk group stratification using pretreatment CT scans in HNSCC.

**Materials and Methods:** We analyzed data of 2485 patients from three retrospective HNSCC cohorts (RADCURE, HN1, HN-PET-CT), treated between 2004 and 2017 with available pre-treatment CTs and primary gross tumor volume (GTVp) segmentations. The RADCURE cohort was split into training (n=1464) and test (N=606), with HN1 (n=131) and HN-PET-CT (n=284) as additional test cohorts. FM-based MIL models (2D, multiview and 3D) for 2-year endpoint prediction and risk stratification wre evaluated based on area under the receiver operator curve (AUROC) and Kaplan-Meier (KM) with hazard ratios (HR), compared with radiomics and assessed for multimodal enhancement with clinical baselines.

**Results:** 2D MIL models achieved 2-year test AUROCs of 0.75-0.84 (OS), 0.66-0.75 (LRC) and 0.71-0.78 (FFDM), outperforming multiview and 3D MIL (AUROCs: 0.50-0.77, p≥0.15) and comparable or superior to radiomics (AUROCs: 0.64-0.74, p≥0.012). Significant stratification was observed (HRs: 2.14-4.77, p≤0.039). Multimodal enhancement of 2-year OS/FFDM (AUROCs: 0.82-0.87, p≤0.018) was observed for patients without human papilloma virus positive (HPV+) tumors.

**Conclusion:** FM-based MIL demonstrates promise in HNSCC risk prediction, showing similar or superior performance to radiomics and enhancing clinical baselines in non-HPV+ patients.

**Key Results:**

- First end-to-end study using both foundation models and multiple instance learning for outcome prediction in head and neck squamous cell carcinoma.
- Multiple instance learning approaches predict clinically-relevant 2-year endpoints and stratify patients across external cohorts with similar or better performance than handcrafted radiomics.
- Multimodal inclusion of clinical and multiple instance learning information improve clinical baseline models in patients without human papillomavirus positive tumors.

## Introduction

Despite multimodal treatment, OS at 5-years remains at 30-70% for patients with HNSCC with existing prognostic methods based on the TNM staging system(1). While HPV infection status is the only validated biochemical test for HNSCC, with HPV16+ oropharyngeal tumors being more radiosensitive and presenting better response to treatment(2), there is no current consensus for its integration into treatment planning. Factors such as extranodal extension have been shown as having a negative prognostic effect but are not consistently quantified in a unified way(3). This highlights the need for more precise prognostic biomarkers that can stratify patients into risk groups and further enable patient-tailored treatment(4) such as dose de-escalation treatment for radiotherapy to reduce patient toxicity in low-risk patients(5) or detection of patients with persistent tumors to consider for second-line treatment(6).

CT scans are routinely acquired for every patient with HNSCC and have been proposed as sources to extract quantitative, non-invasive biomarkers from. Radiomics was initially devised to extract handcrafted features from radiological images to create such image-based biomarkers(7). Despite large amounts of studies and efforts for standardization(8), adoption of such handcrafted radiomics biomarkers in clinical settings remains limited. One of the main drawbacks of such studies is the time-intensive requirement for manual image annotations and the variability across studies attempting to predict the same outcome(9). In contrast, Deep Learning (DL) approaches offer flexibility by learning image representations without the need for prior handcrafted feature definition. DL methods have been developed for clinical applications in HNSCC to predict LRC from PET/CT images(10), epithelial growth factor receptor mutation status(11) or HPV infection in oropharyngeal cancer patients(12) in CT. However, DL methods require larger datasets for training models than handcrafted radiomics studies, which limits the implementation of such methods in practice.

An emerging area of AI research in medicine is FMs, models trained on large and unlabeled datasets to capture general patterns in the data. Then, FMs can act as backbones on downstream tasks, bypassing the need to train models from scratch. This approach allows for the extraction of general-purpose features and the development of predictive DL models from smaller datasets. Currently available FMs for extracting features that utilize radiological images for training include BioMedClip(13), using 2D images of a single view or the FM by Pai et al.(14) using 3D bounding boxes. Some of the current FMs require aggregation methods to analyze CT scans. Multiple instance learning (MIL) is a weakly-supervised paradigm(15) that can serve as a solution by creating groups of data instances from each patient(16). MIL allows for the simultaneous learning of relationships between data instances, such as different 2D slices of a CT image, and a target, such as the patient’s outcome. This is potentially advantageous as it enables handling data with ambiguous labels or identification of relevant patterns when extensive labeling of specific regions or pixels is impractical. DL-based MIL has been successfully employed within the histopathology domain for tasks such as microsatellite instability classification in colorectal cancer(17) or immunotherapy response on solid tumors(18). In the radiological domain, MIL has been explored(19,20) but has not been employed before for outcome prediction using pretreatment CT images in HNSCC with FMs features.

The present study aims to implement, for the first time, DL-based MIL approaches applied to FM features from pretreatment CTs of HNSCC patients for outcome prediction. As we hypothesize that MIL can aggregate FM features to learn prognostic biomarkers from the CT images, we conduct a comprehensive study including different FMs as feature extractors combined with MIL approaches to predict three different endpoints for HNSCC. The results of this study are evaluated across three different test cohorts and compared with handcrafted radiomics models and a clinical baseline. Finally, we explore correlations with clinical factors as well as the enhancement our models provide to the clinical baseline for prognosis in the overall patient population as well as non-HPV+ HNSCC patients.

## Materials and Methods

### Patient cohorts and endpoint dichotomization

This retrospective study considers three cohorts with already-deidentified patients extracted from The Cancer Imaging Archive(21): the RADCURE cohort(22), the HN-PET-CT cohort(23,24), and the HN1 cohort(7,25). The RADCURE cohort consists of 3346 head and neck cancer patients treated with definitive radiotherapy or radiochemotherapy between 2005 and 2017 at the Princess Margaret Cancer Centre, Toronto, Canada. The HN-PET-CT cohort consists of 298 HNSCC patients from 4 different hospitals in Canada. These patients had received either definitive radiotherapy, radiochemotherapy or a combination of surgery and either adjuvant radiotherapy or radiochemotherapy between 2006 and 2014. The HN1 cohort consists of 137 HNSCC patients treated with radiotherapy or radiotherapy with concurrent chemotherapy at the MAASTRO clinic, Maastricht, the Netherlands starting from 2004. Patient consent was deemed unnecessary due to the public nature of the datasets.

Inclusion and exclusion criteria for the study are summarized in Figure 1. Patients who underwent surgery, had metastases upon diagnosis, or lacked proof of presenting with HNSCC through either histological proof or mention in their source publications were excluded. Inclusion criteria were defined as the availability of pretreatment CTs and GTVp segmentations. Endpoints were OS, LRC, and FFDM dichotomized at the 2-year mark similar to a prior study(26) for patient retention purposes. For endpoint dichotomization, patients who were lost to follow-up before the 2-year mark or had ambiguous endpoint data, such as presenting recurrence the same day of treatment, were dropped for that endpoint. Patients with persisting disease were excluded from the 2-year LRC endpoint. OS was defined as the time from treatment start to death by any cause or loss to follow-up. LRC was defined from treatment start to local or regional tumor recurrence or loss to follow-up. FFDM was defined from treatment start until detection of distant metastasis or loss to follow-up.

**Figure 1.**
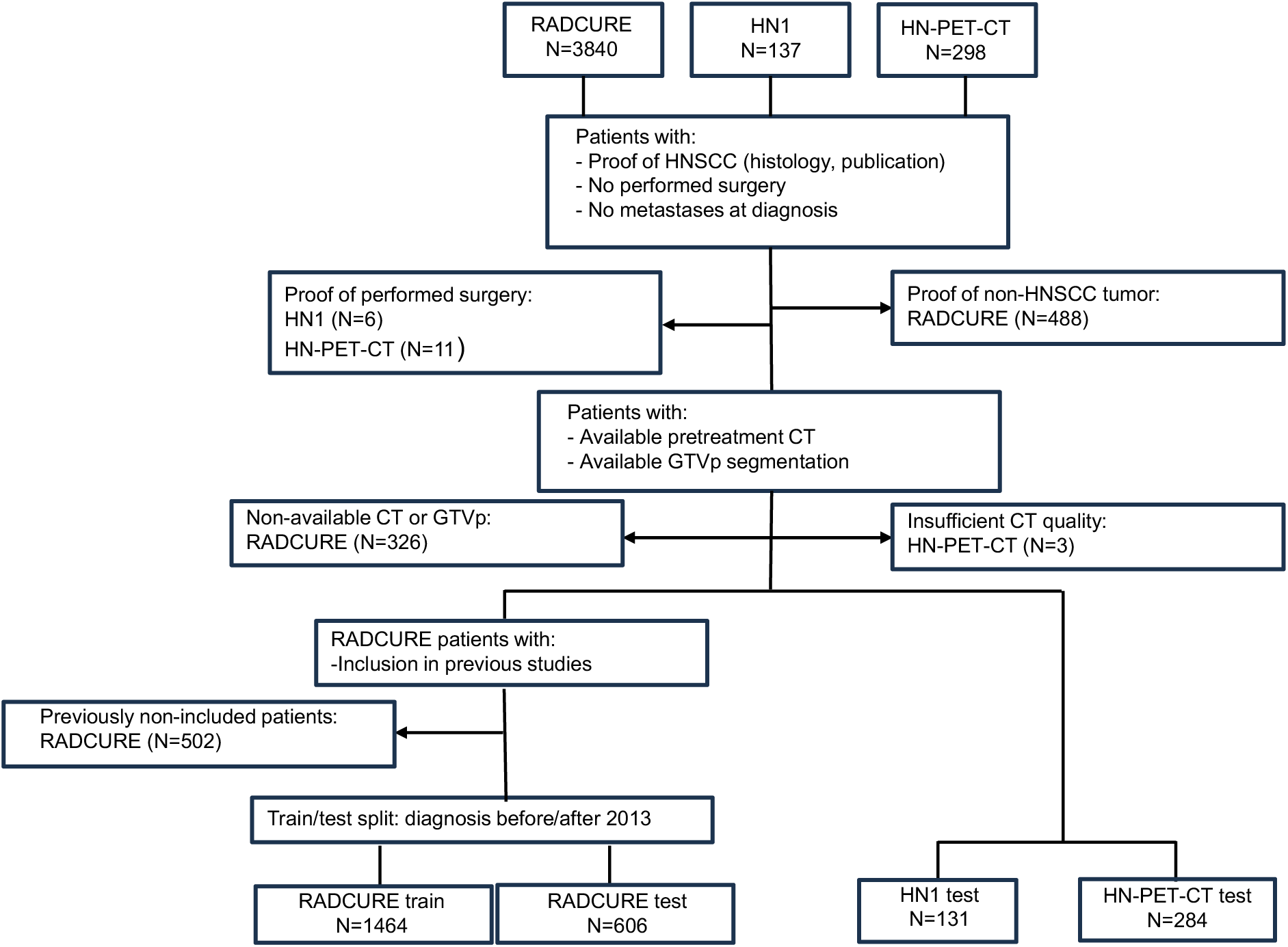
Flowchart showing inclusion and exclusion criteria for study cohorts as well as cohorts used for training models and for testing. Excluded patients are shown per cohort and the reason for the exclusion.

Included patients were reported in previous studies (7,24,26) dealing with outcome prediction for OS (7,26) or LRC and FFDM (24), with either non-MIL DL or handcrafted-radiomics approaches. In contrast, the current study deals with the application of FMs as feature extractors for MIL modeling of 2-year endpoint prediction, stratification, and multimodal integration alongside clinical variables for OS, LRC and FFDM in HNSCC patients.

### Study design

Within this retrospectively-designed study cohorts were divided into training and test cohorts to discern whether 2D, multiview or 3D FM features are best for HNSCC outcome prediction with MIL, if MIL models outperform handcrafted radiomics features and whether MIL scores add value to already-available clinical parameters (Fig. 2A). In order to obtain features from the CT images to be used in MIL modeling(Fig. 2B), two approaches were considered for feature extraction: a 2D approach to extract information (Fig. 2C) from the three different views of the CT image (axial, coronal and sagittal) and a 3D approach based on sampling volumes within the CT (Fig. 2D).

**Figure 2.**
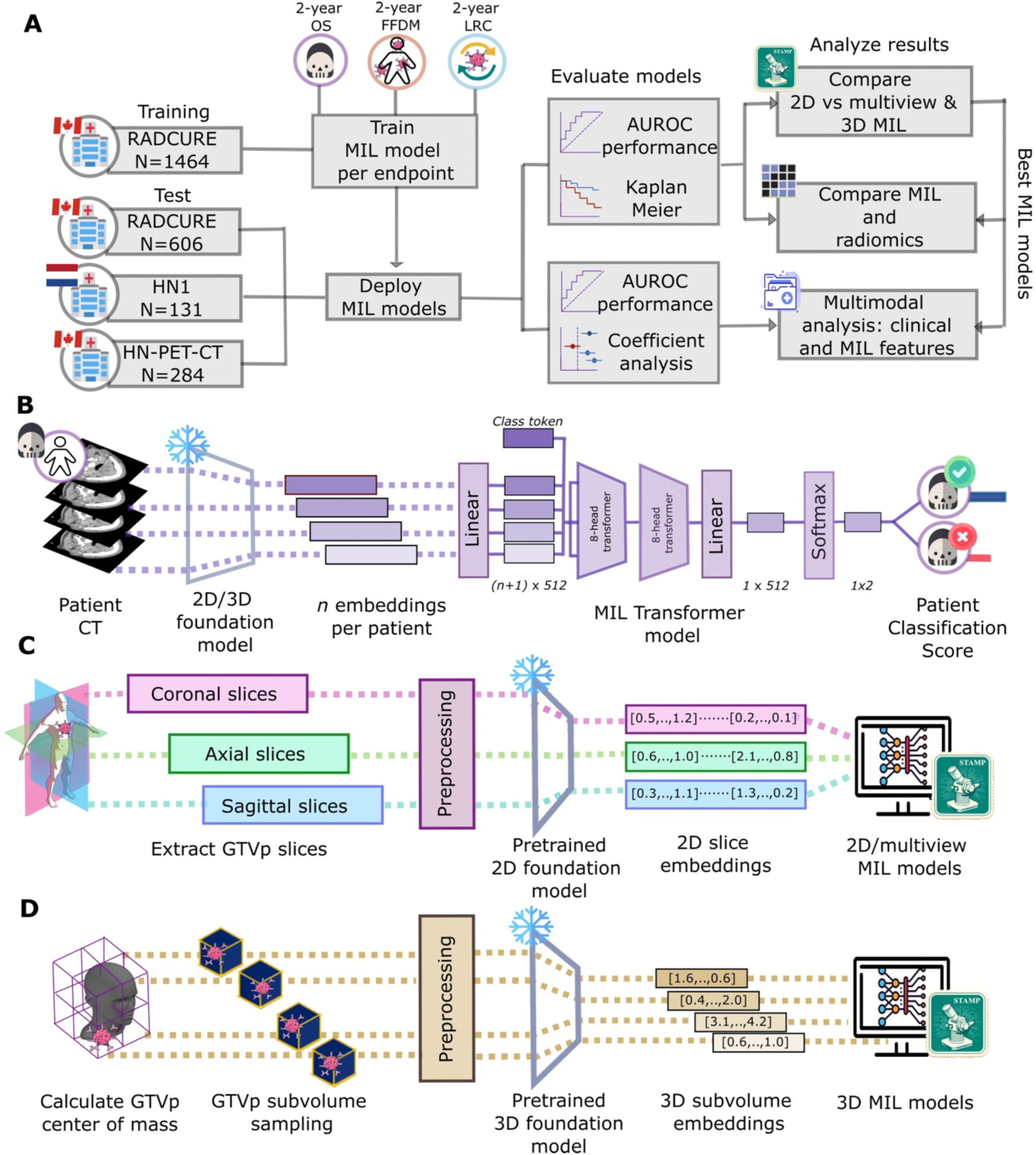
Workflow for overall study design, modeling and extraction. MIL models were trained to predict one of three 2-year endpoints on the training cohort and evaluated on three test cohorts (A) for each of three 2-year endpoints. For 2-year OS (B), for example, a patient would have embeddings extracted from the CT which would be used as input into a MIL transformer, learning simultaneously on embeddings from all CTs slices/subvolumes and pooling them to return patient-level scores for 2-year OS. Embeddings are extracted from FMs using either (C) axial or multiple views of CT slices containing the GTVp or from (D) subvolumes sampled around the GTVp center for a 3D approach.

### Preprocessing and feature extraction

For all CT images, the patient body was segmented through TotalSegmentator(27) and used to mask out the patient bed. For 2D MIL, for each CT view all 2D CT slices containing the GTVp mask were selected. CT images and GTVp segmentations were extracted and resampled to voxel spacing of 1×1×1 mm³ through the SimpleITK Python library. Hounsfield Units (HU) were clipped with a soft tissue windowing approach with a level of 50 HU and a window width of 120 HU. Images were converted into grayscale, cropped on the GTVp center of mass to size 224×224 pixels and normalized to FM preprocessing parameters (Supplementary Table 1). Different pretrained feature extractors were considered to obtain slice-level representations of the patient: i) ImageNet-pretrained ResNet50, ii) ImageNet-pretrained SwinViT(28), and iii) the PMC-15 pretrained vision encoder extracted from the BioMedClip FM(13). 2D models were constructed using either axial view features or features from all three CT views (multiview models).

In 3D MIL, for the FM by Pai et al.(14) images were preprocessed according to FM specifications of the original publication: values normalized to a range of −1024 and 2048 HU and resampled to 1×1×1 mm³. To capture different contexts of the tumor and its surrounding tissue, 50×50×50 voxel volumes were created by randomly sampling 100 times from a multivariate normal distribution with variance of 16 and with the coordinates of the center of mass of the GTVp as mean. An ImageNet-pretrained inflated 3D ResNet50(29) was also considered, with features extracted similarly to the previously mentioned FM and using the same HU windowing.

### MIL model training

Extracted features were input into a MIL Transformer (30,31) using the STAMP package version 1.0.1. A MIL model was trained for each endpoint through class-stratified 5-fold cross-validation. For each MIL model and endpoint, the model with the area under the receiver operator curve (AUROC) closest to the median AUROC value of the cross-validation holdout set was chosen for deployment in the test cohorts for classification and stratification purposes. Subgroup analysis to assess performance changes based on contrast enhancement status were also conducted. Training specifications are shown in the Supplementary Materials.

### Handcrafted radiomics extraction and modeling

Morphological, statistical, and texture features were extracted alongside from the base image using PyRadiomics with only IBSI-compliant features. Feature selection consisted of variance filtering, z-score standardization, hierarchical clustering with correlation distance cutoff of 0.20 for cluster membership with average Spearman correlation for cluster representative selection followed by minimum redundancy maximum relevance selection similar to another study for HNSCC handcrafted radiomics modelling(32). The number of features selected was the number of samples in the minority class divided by 10. Selected features were then input into a logistic regression model for training and tested on the test cohorts. Subgroup analysis to assess performance changes based on contrast enhancement status were also conducted. Extraction parameters are shown in Supplementary Table 2.

### Clinical baseline and multimodal enhancement

Baseline clinical models were developed using the following factors: T stage, N stage, HPV status, age, sex and treatment using a logistic regression. Relationships between MIL scores and clinical baseline features were assessed by the coefficients of a multivariate logistic regression model containing the MIL scores and the clinical baseline features on the training cohort as well as with a correlation analysis between MIL scores and clinical baseline factors in the external test cohorts. Enhancement of model performance by inclusion of MIL scores was assessed in all patients of the test cohorts as well as in the subgroup of patients without HPV+ tumors.

### Statistical analysis

Statistical comparisons between categorical variables in the training and test cohorts were conducted through the chi-squared test or the exact Fisher test. Continuous variables were compared through the Mann-Whitney U-test. The AUROC was chosen as a summary metric of classification performance for 2-year endpoints. AUROC 95% confidence intervals (95% CI) were obtained from the empirical distribution through 1000 class-stratified bootstraps. Risk group stratification was conducted through the KM estimator using the 2-year MIL scores as risk scores. The cutoff was the median score on the training cohort, and risk group separation was assessed through the Cox HR and the logrank test p-value. For explainability, attention scores were generated and the top-3 CT slices from selected patients were displayed with element-wise gradcam used to visualize the regions important for model-prediction within the slice through the gradcam Python library.

AUROC differences were assessed through score permutation tests: for 1000 bootstraps, predictions from randomly selected patients were permuted between models and the resulting AUROCs were used to define a null distribution and heuristic p-value. All statistical tests were conducted in a two-sided manner unless specified. Significance of MIL scores in multivariate logistic regression was assessed through significance of log-odds ratios (LOR). Correlations of clinical baseline factors with MIL scores were assessed through Spearman rho or through point-biserial correlation. A p-value threshold of 0.05 was chosen for significance. Multiple comparison adjustment was performed through the Benjamini-Hochberg method. All analyses were conducted in Python 3.11 or R version 4.4.0. Code available at: https://github.com/KatherLab/HNSCC_FM_MIL_study.

## Results

### Patient cohort clinical characteristics

The characteristics of the patients in the RADCURE train (N=1404), RADCURE test (N=606), HN1 (N=131), and HN-PET-CT (N=284) cohorts are shown in Table 1. The RADCURE test cohort patients had significantly lower ECOG stages, higher N-stages, and a higher proportion of oropharyngeal tumors (all p<0.001). The HN1 cohort had a significantly higher proportion of patients with recurrence before 2 years, higher ECOG status, and significantly fewer HPV+ patients (all p<0.001). Tumors in the HN-PET-CT cohort had significantly larger volume, a higher proportion of patients treated with chemotherapy and significantly higher 2-year OS (all p<0.001). CT scanner parameters across cohorts are shown in Supplementary Table 3. No statistically significant differences in age, sex, radiation dose, number of fractions, and 2-year FFDM were observed across cohorts.

**Table 1.** Demographic, clinical and endpoint data for included patients of the training and test cohorts. Continuous variables are indicated by median [interquartile range]. Categorical variables are indicated with their number and percentage per category. Statical comparisons between training and each test cohort are also shown. Missing categories/variables within one of the cohorts are indicated by NA. T stage, N stage and Staging were acquired following the American Joint Committee on Cancer 7th Edition criteria. Number of patients dropped for each 2-year dichotomised prognostic endpoint are labelled as Dropout.

|  |  | **RADCURE**<br>**train**<br>**(n=1464)** | **RADCURE**<br>**test**<br>**(n=606)** | **p-value** | **HN1**<br>**(n=131)** | **p-value** | **HN-PET-CT**<br>**(n=284)** | **p-value** |
| --- | --- | --- | --- | --- | --- | --- | --- | --- |
| **Age (years)** |  | 63.20<br>[55.70-71.60] | 63.65<br>[57.20-70.10] | 0.76 | 62.00<br>[56.00-67.50] | 0.10 | 63.00<br>[57.00-70.25] | 0.86 |
| **Total Dose (Gy)** |  | 70.00<br>[64.00-70.00] | 70.00<br>[70.00-70.00] | 1.00 | 68.00<br>[68.00-70.00] | 1.00 | NA | NA |
| **Dose fractions**<br>**(# fractions)** |  | 35<br>[33-35] | 35<br>[35-35] | 1.00 | 34<br>[34-35] | 1.00 | NA | NA |
| **GTVp (cm<sup>3</sup>)** |  | 14.55<br>[5.00-30.52] | 13.68<br>[4.87-30.14] | 0.66 | 14.15<br>[3.69-33.71] | 0.46 | 23.72<br>[11.58-40.55] | **<0.001** |
| **Gender** | Male | 1200 (82.0) | 501 (82.7) | 1.00 | 106 (80.9) | 0.86 | 219 (77.1) | 0.067 |
|  | Female | 264 (18.0) | 105 (17.3) |  | 25 (19.1) |  | 65 (22.9) |  |
| **ECOG Status** | ECOG-0 | 878 (60.0) | 335 (55.3) | **<0.001** | 60 (49.6) | **<0.001** | NA | NA |
|  | ECOG-1 | 408 (27.9) | 248 (40.9) |  | 57 (47.1) |  | NA |  |
|  | ECOG-2 | 131 (9.0) | 20 (3.3) |  | 3 (2.5) |  | NA |  |
|  | ECOG-3 | 27 (1.8) | 3 (0.5) |  | 1 (0.8) |  | NA |  |
|  | ECOG-4 | 5 (1.0) | 0 (0.0) |  | 0 (0.0) |  | NA |  |
|  | Unknown | 15 (0.3) | 0 (0.0) |  | 0 (0.0) |  | NA |  |
| **Site** | Oral Cavity | 70 (4.8) | 10 (1.7) | **<0.001** | 0 (0.0) | **<0.001** | 0 (0.0) | **<0.001** |
|  | Pharynx | 0 (0.0) | 0 (0.0) |  | 0 (0.0) |  | 0 (0.0) |  |
|  | Hypopharynx | 110 (7.5) | 28 (4.6) |  | 0 (0.0) |  | 13 (4.6) |  |
|  | Larynx | 534 (36.5) | 172 (28.4) |  | 0 (0.0) |  | 45 (15.8) |  |
|  | Nasal Cavity | 27 (1.8) | 18 (3.0) |  | 46 (35.1) |  | 0 (0.0) |  |
|  | Oropharynx | 700 (47.8) | 362 (59.7) |  | 85 (64.9) |  | 190 (66.9) |  |
|  | Other | 23 (1.6) | 16 (2.6) |  | 0 (0.0) |  | 28 (9.9) |  |
|  | Unknown | 0 (0.0) | 0 (0.0) |  | 0 (0.0) |  | 8 (6.1) |  |
| **T stage** | T1 | 292 (20.0) | 119 (19.6) | 0.39 | 33 (25.2) | **<0.001** | 36 (12.7) | **0.02** |
|  | T2 | 453 (30.9) | 210 (34.7) |  | 32 (24.4) |  | 101 (35.6) |  |
|  | T3 | 425 (29.0) | 160 (26.4) |  | 23 (17.6) |  | 92 (32.4) |  |
|  | T4 | 268 (18.3) | 107 (17.7) |  | 43 (32.8) |  | 46 (16.2) |  |
|  | Unknown | 26 (1.8) | 10 (1.6) |  | 0 (0.0) |  | 9 (3.2) |  |
| **N stage** | N0 | 621 (42.4) | 203 (33.5) | **0.002** | 58 (44.3) | 0.25 | 59 (20.8) | **<0.001** |
|  | N1 | 109 (7.5) | 55 (9.1) |  | 16 (12.2) |  | 38 (13.4) |  |
|  | N2 | 672 (45.9) | 322 (53.1) |  | 54 (41.2) |  | 164 (57.7) |  |
|  | N3 | 62 (4.2) | 26 (4.3) |  | 3 (2.3) |  | 19 (6.7) |  |
|  | Unknown | 0 (0.0) | 0 (0.0) |  | 0 (0.0) |  | 0 (0.0) |  |
| **Staging** | I | 223 (15.2) | 69 (11.4) | **0.005** | 24 (18.3) | 0.39 | NA | NA |
|  | II | 194 (13.3) | 78 (12.9) |  | 11 (8.4) |  | NA |  |
|  | III | 236 (16.1) | 75 (12.4) |  | 22 (16.8) |  | NA |  |
|  | IV | 810 (55.3) | 381 (62.9) |  | 74 (56.4) |  | NA |  |
|  | Unknown | 1 (0.0) | 3 (0.4) |  | 0 (0.0) |  | NA |  |
| **HPV16** | Positive | 459 (31.4) | 290 (47.9) | 0.31 | 22 (16.8) | **<0.001** | 71 (25.0) | 0.32 |
|  | Negative | 232 (15.8) | 127 (21.0) |  | 56 (42.7) |  | 45 (15.8) |  |
|  | Unknown | 773 (52.8) | 189 (31.2) |  | 53 (40.5) |  | 168 (59.2) |  |
| **Contrast** | Yes | 771 (52.7) | 521 (86.0) | **<0.001** | 0 (0.0) | **<0.001** | 0 (0.0) | **<0.001** |
|  | No | 693 (47.3) | 85 (14.0) |  | 131 (100.0) |  | 284 (100.0) |  |
| **Chemotherapy** | Yes | 976 (66.7) | 354 (58.4) | **<0.001** | 34 (26.0) | **<0.001** | 236 (83.0) | **<0.001** |
|  | No | 488 (33.3) | 252 (41.6) |  | 97 (74.0) |  | 48 (17.0) |  |
| **2-year LRC** | Recurrent | 159 (10.9) | 69 (11.4) | 0.14 | 27 (20.6) | **0.003** | 29 (10.2) | 0.50 |
|  | Non-recurrent | 1063 (72.6) | 361 (59.6) |  | 88 (67.1) |  | 229 (80.6) |  |
|  | Dropout | 242 (16.5) | 176 (29.0) |  | 16 (12.2) |  | 26 (9.2) |  |
| **2-year OS** | Dead | 294 (20.1) | 89 (14.7) | 0.38 | 30 (22.9) | 0.52 | 26 (9.2) | **<0.001** |
|  | Alive | 1167 (79.7) | 401 (66.2) |  | 101 (77.1) |  | 241 (84.9) |  |
|  | Dropout | 3 (0.2) | 116 (19.1) |  | 0 (0.0) |  | 17 (5.9) |  |
| **2-year FFDM** | Metastatic | 147 (10.0) | 61 (10.1) | 0.22 | 7 (5.3) | 0.17 | 29 (10.2) | 0.90 |
|  | Non-metastatic | 1098 (75.0) | 367 (60.6) |  | 96 (73.3) |  | 228 (80.3) |  |
|  | Dropout | 219 (15.0) | 178 (29.3) |  | 28 (21.2) |  | 27 (9.5) |  |

### Performance of the clinical baseline model

First, we evaluated the predictive performance of the clinical baseline models (Suppl. Tables 4-6). For 2-year OS the model showed AUROCs of 0.83 [0.78-0.88], 0.82 [0.73-0.89] and 0.78 [0.70-0.86] for the RADCURE test, HN1 and HN-PET-CT cohorts. For 2-year LRC, the model showed AUROCs of 0.81 [0.76-0.86], 0.79 [0.69-0.86] and 0.74 [0.66-0.82] for RADCURE test, HN1 and HN-PET-CT respectively. Finally, the model showed AUROCs of 0.76 [0.70.0.82], 0.86 [0.78-0.93] and 0.74 [0.66-0.81] for the RADCURE test, HN1 and HN-PET-CT cohorts for 2-year FFDM.

### Performance of the handcrafted radiomics model

Secondly, we assessed the performance of the handcrafted radiomics models (Suppl. Tables 7-9). For 2-year OS the AUROCs were of 0.73 [0.68-0.80], 0.73 [0.64-0.85] and 0.67 [0.56-0.79] for the RADCURE-test, HN1 and HN-PET-CT cohorts. For 2-year LRC, AUROCs were 0.68 [0.62-0.76] for RADCURE-test, 0.73 [0.60-0.84] for HN1 and 0.63 [0.52-0.74] for HN-PET-CT. For 2-year FFDM, AUROCs were 0.71 [0.64-0.77], 0.68 [0.52-0.83] and 0.67 [0.56-0.79] for RADCURE test, HN1 and HN-PET-CT respectively. Models trained using the subgroups of non-CE (Suppl. Tables 10-12) and CE patients (Suppl. Tables 13-15) did not show significantly higher performance (p≥0.20) when compared to the models trained on the entire training cohort (Suppl. Table 16).

### Deep learning models classify 2-year events for OS, LRC and FFDM

For MIL models, performances for the best models per model type and endpoint are shown in Figure 3 alongside handcrafted radiomics and clinical models. Multiview and 3D models’ performances were not significantly higher than 2D MIL models with axial features (p≥0.15, Suppl. Table 17). For MIL models with 2D axial features, the best model for 2-year OS showed AUROCs of 0.75 [0.69-0.80], 0.77 [0.68-0.85] and 0.84 [0.77-0.90] for the RADCURE test, HN1 and HN-PET-CT cohorts respectively. For 2-year LRC, AUROCs of 0.75 [0.68-0.81], 0.66 [0.52-0.78] and 0.72 [0.61-0.80] were obtained for the RADCURE test, HN1 and HN-PET-CT cohorts respectively. Finally, for 2-year FFDM, the best-performing model achieved AUROCs of 0.78 [0.71-0.84], 0.77 [0.58-0.93] and 0.73 [0.61-0.83] for each test cohort. ROC curves across all extractors and endpoints are shown in Suppl. Fig. 1-3. Contrast subgroup-specific models did not significantly outperform models trained on the entire training cohort (p≥0.20, Suppl.Table 18). When compared against handcrafted radiomics models as shown in Table 2, 2D MIL models showed similar (p≥0.15) performances except for 2-year OS in the HN-PET-CT cohort where 2D MIL significantly outperformed handcrafted radiomics (p=0.012).

**Figure 3.**
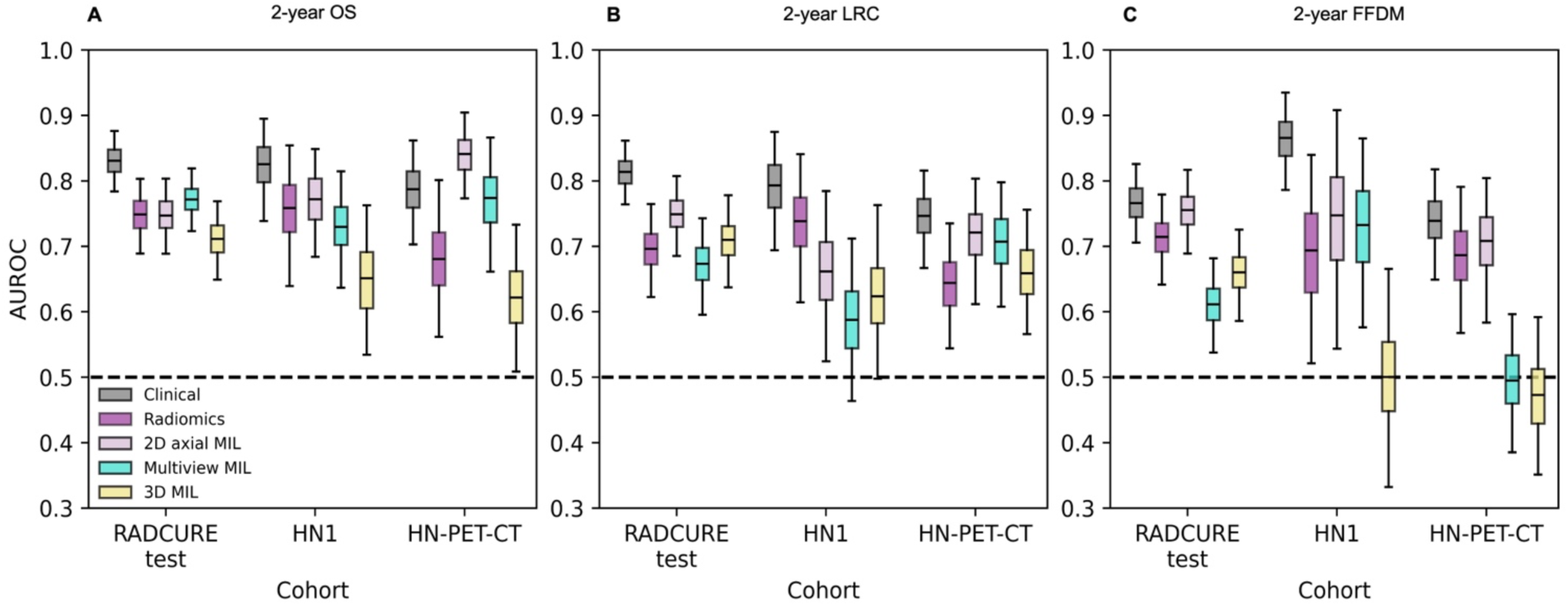
Performance summary for 2-year endpoint classification for the clinical baseline, handcrafted radiomics and the best 2D axial, multiview and 3D MIL models shown across the three test cohorts. AUROC distributions from 1000 class-stratified bootstraps are shown for (A) 2-year OS, (B) 2-year LRC and (C) 2-year FFDM.

**Table 2.** AUROC across 2-year endpoints and cohort for the obtained radiomics models alongside p-value of one-sided permutation tests with alternative hypothesis of superiority of MIL models over radiomics models.

| Endpoint | Cohort | AUROC [95% CI]<br>2D MIL | AUROC [95% CI]<br>radiomics | p-value |
| --- | --- | --- | --- | --- |
| 2-year OS | RADCURE test | 0.75 [0.69-0.80] | 0.74 [0.68-0.80] | 0.50 |
| 2-year OS | HN1 | 0.77 [0.68-0.85] | 0.74 [0.64-0.85] | 0.50 |
| 2-year OS | HN-PET-CT | 0.84 [0.77-0.90] | 0.68 [0.56-0.79] | **0.012** |
| 2-year LRC | RADCURE test | 0.75 [0.68-0.81] | 0.69 [0.62-0.76] | 0.15 |
| 2-year LRC | HN1 | 0.66 [0.52-0.78] | 0.74 [0.61-0.84] | 0.84 |
| 2-year LRC | HN-PET-CT | 0.72 [0.61-0.80] | 0.64 [0.53-0.74] | 0.15 |
| 2-year FFDM | RADCURE test | 0.78 [0.71-0.84] | 0.71 [0.64-0.77] | 0.21 |
| 2-year FFDM | HN1 | 0.75 [0.55-0.92] | 0.68 [0.52-0.83] | 0.48 |
| 2-year FFDM | HN-PET-CT | 0.71 [0.59-0.80] | 0.67 [0.66-0.79] | 0.53 |

### Deep learning MIL scores stratify patients for time-to-event analysis

Next, we investigated whether MIL models could stratify the full time-to-event endpoints. For the best 2D axial MIL models, OS showed significant stratification (p<0.001) across all three test cohorts as shown in Figure 4. For LRC and FFDM (Suppl. Fig. 4), the 2D axial MIL models were able to significantly stratify all (p≤0.039) and two out of three cohorts (p≤0.032), respectively, tending to show higher HRs and more significant stratification when compared to handcrafted-radiomics (p≤0.20) for OS but not higher than clinical baselines as seen in Table 3. Multiview and 3D models did not show better stratification than 2D MIL models (Suppl. Fig. 5-6). KM curves for clinical baseline and handcrafted radiomics models are shown in Suppl. Fig. 7-8.

**Figure 4.**
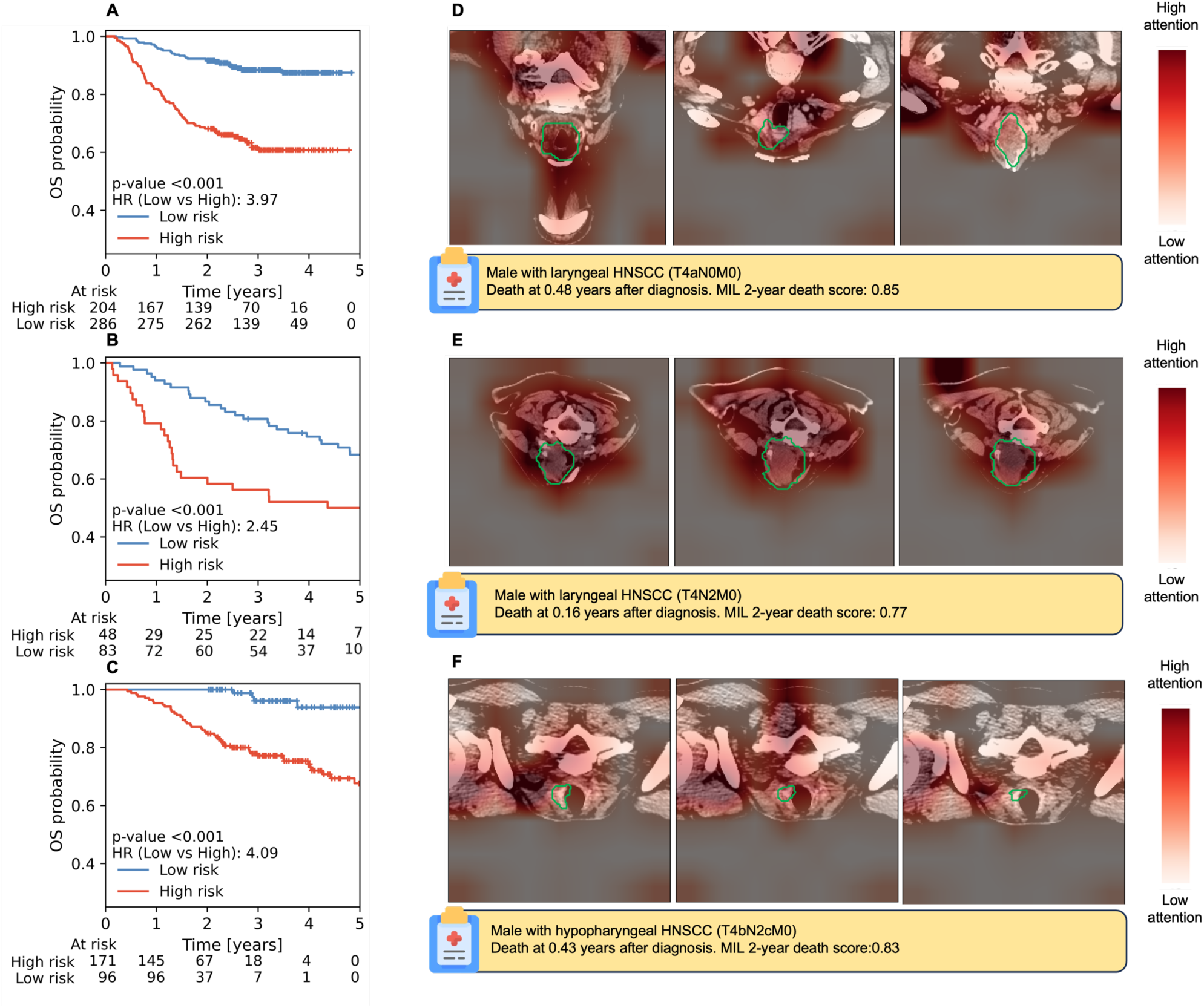
KM plots for OS stratification with the best 2D MIL model for the (A) RADCURE test, (B) HN1 and (C) HN-PET-CT cohorts alongside exemplary axial CT slices with heatmaps for the three CT slices with highest attention belonging to a high-risk patient for RADCURE test (D), HN1 (E) and HN-PET-CT (F) are shown alongside patient characteristics. CT scans are shown after image preprocessing and with a green contour indicating the GTVp segmentation.

**Table 3.**
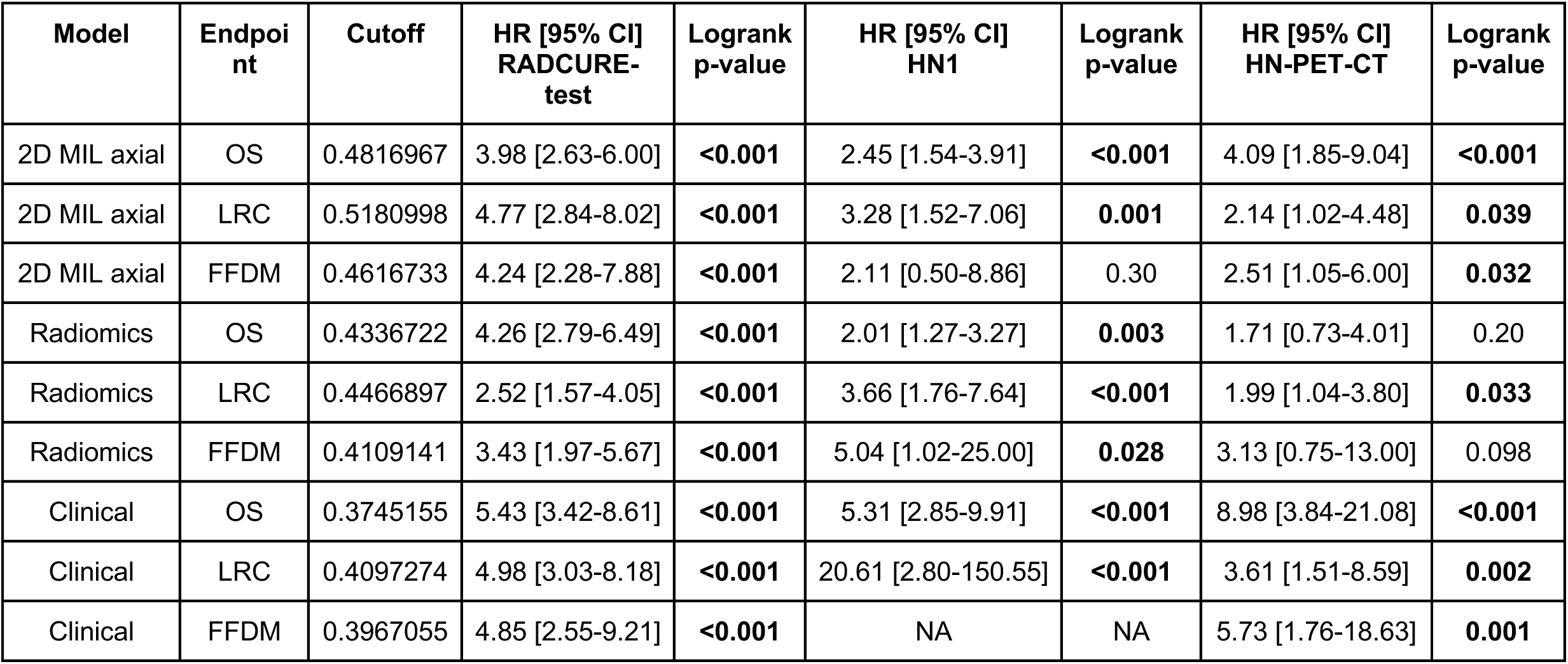
KM stratification summary with HR [95% CI] between high and low-risk groups across all test cohorts for 2D axial MIL, radiomics and clinical models alongside logrank p-value and cutoff extracted from the training cohort. NAs indicate that stratification into two groups produced only one group with events and was thus not possible to assess separation. HRs smaller than 1 indicate that patients with higher risk scores have a smaller probability of event.

| Model | Endpoint | Cutoff | HR [95% CI]<br>RADCURE-test | Logrank<br>p-value | HR [95% CI]<br>HN1 | Logrank<br>p-value | HR [95% CI]<br>HN-PET-CT | Logrank<br>p-value |
| --- | --- | --- | --- | --- | --- | --- | --- | --- |
| 2D MIL axial | OS | 0.4816967 | 3.98 [2.63-6.00] | <0.001 | 2.45 [1.54-3.91] | <0.001 | 4.09 [1.85-9.04] | <0.001 |
| 2D MIL axial | LRC | 0.5180998 | 4.77 [2.84-8.02] | <0.001 | 3.28 [1.52-7.06] | 0.001 | 2.14 [1.02-4.48] | 0.039 |
| 2D MIL axial | FFDM | 0.4616733 | 4.24 [2.28-7.88] | <0.001 | 2.11 [0.50-8.86] | 0.30 | 2.51 [1.05-6.00] | 0.032 |
| Radiomics | OS | 0.4336722 | 4.26 [2.79-6.49] | <0.001 | 2.01 [1.27-3.27] | 0.003 | 1.71 [0.73-4.01] | 0.20 |
| Radiomics | LRC | 0.4466897 | 2.52 [1.57-4.05] | <0.001 | 3.66 [1.76-7.64] | <0.001 | 1.99 [1.04-3.80] | 0.033 |
| Radiomics | FFDM | 0.4109141 | 3.43 [1.97-5.67] | <0.001 | 5.04 [1.02-25.00] | 0.028 | 3.13 [0.75-13.00] | 0.098 |
| Clinical | OS | 0.3745155 | 5.43 [3.42-8.61] | <0.001 | 5.31 [2.85-9.91] | <0.001 | 8.98 [3.84-21.08] | <0.001 |
| Clinical | LRC | 0.4097274 | 4.98 [3.03-8.18] | <0.001 | 20.61 [2.80-150.55] | <0.001 | 3.61 [1.51-8.59] | 0.002 |
| Clinical | FFDM | 0.3967055 | 4.85 [2.55-9.21] | <0.001 | NA | NA | 5.73 [1.76-18.63] | 0.001 |

### Deep learning MIL scores are significant predictors in multivariate analysis

When adjusted for clinical baseline factors in a multivariate setting (Fig. 5A-C) 2D MIL scores remained significant predictors (p≤0.001), with increases in MIL scores resulting in higher probability of presenting 2-year events for all endpoints (Suppl. Tables 19-21) For 2-year OS, the 2D MIL clinically-adjusted LOR was 0.98 [0.82-1.05], for 2-year LRC LOR was 0.89 [0.77-1.01] and for 2-year FFDM LOR was 0.72 [0.56-0.87]. When examining correlations with clinical factors in the external test cohorts (Fig. 5D-F), MIL scores consistently showed mild to strong correlation with higher T stages for 2-year OS (rho:0.26-0.55), 2-year LRC (rho:0.22-0.51) and 2-year FFDM (rho:0.22-0.65) and consistently showed mild to moderate correlation with N stage for 2-year FFDM (rho:0.32-0.56).

**Figure 5.**
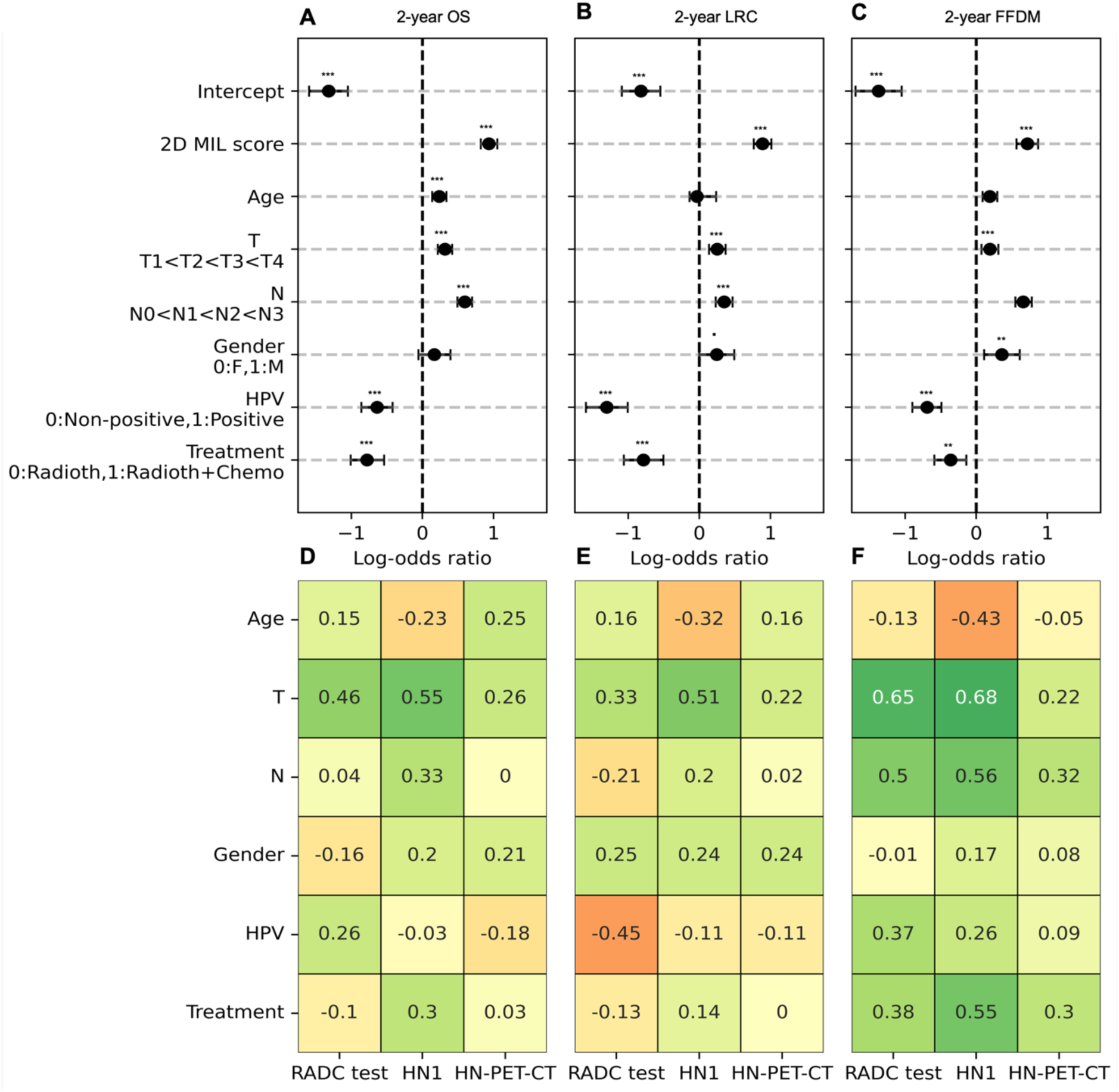
Forest plots for multivariate logistic regression with 2D MIL scores in training (A, B,C) and correlation analysis for test cohorts (D,E,F) per endpoint. Coefficients of multivariate logistic regression (log-odds ratio) are shown for 2-year OS (A), 2-year LRC (B) and 2-year FFDM (C) alongside significance level per coefficient and factoring of variables for analysis. Correlation coefficients of clinical factors through Spearman rho (Age, T stage, N stage) and point-biserial correlation (Treatment, HPV status and gender) are shown for all three test cohorts for 2-year OS (D), 2-year LRC (E) and 2-year FFDM (F) with positive coefficients indicating a positive association between ordering of the clinical factors and increasing 2D MIL scores. Significance levels:.: 0.05<p<0.10, *: 0.01<p<0.05, **:0.001<p<0.01, ***: p<0.001.

### Multimodal input improves model performance

Finally, we investigated multimodal performance enhancement through combining the clinical baseline and 2D MIL scores. When tested on the external test sets, as shown in Figure 6, while models combining clinical baseline features and MIL scores tended to display similar performances compared to the clinical baseline model across endpoints, significant performance improvements were observed (Fig. 6A-C) for 2-year OS in the HN-PET-CT cohort (0.88 [0.81-0.93] vs 0.78 [0.70-0.86], p=0.018). For the subgroup analysis in non-HPV+ patients (Fig. 6D-F), significantly higher performances were observed in the HN-PET-CT cohort for 2-year OS (0.87 [0.79-0.92] vs 0.75 [0.63-0.83], p=0.018) and in the RADCURE test cohort for 2-year FFDM (0.82 [0.74-0.87] vs 0.77 [0.69-0.85], p=0.006).

**Figure 6.**
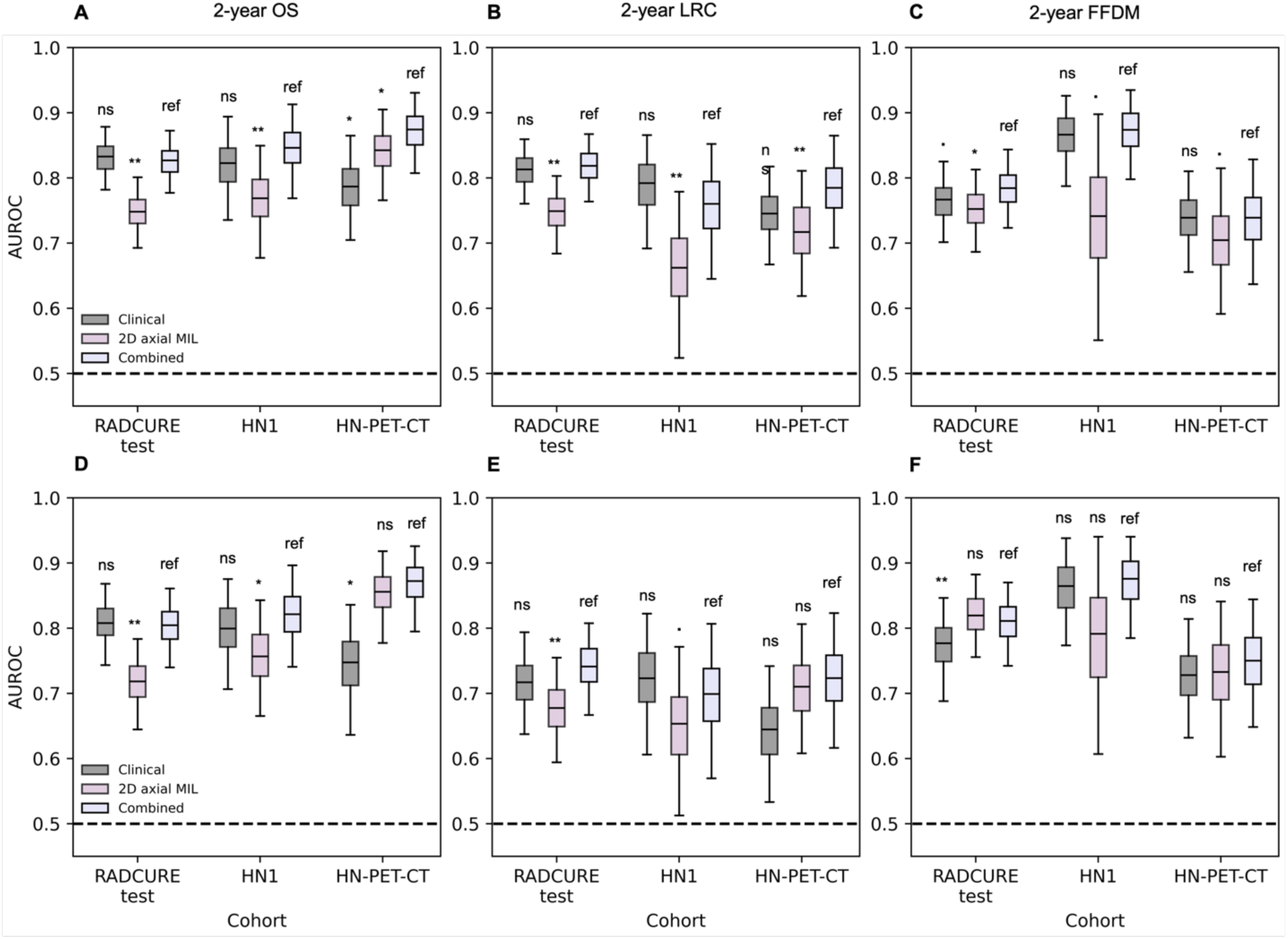
Performance and statistical comparison summary for 2-year endpoint classification for all patients (A,B,C) and patients without HPV-positive HNSCC (D,E,F) in the test cohorts for the clinical baseline, 2D MIL and combined multivariate model with clinical baseline and 2D MIL scores. AUROC distributions from 1000 class-stratified bootstraps are shown across all three test cohorts for (A,D) 2-year OS, (B,E) 2-year LRC and (C,F) 2-year FFDM. Significance levels: ns: non-significant,.:0.05<p<0.10, *: 0.01<p<0.05, **:0.001<p<0.01, ***: p<0.001.

## Discussion

FMs are an emerging area of research for medical AI and have shown great potential as very flexible feature extractors for a variety of downstream tasks in domains such as histopathology(33). Our study assesses, for the first time, FMs as feature extractors with MIL for endpoint prediction in HNSCC using pretreatment CT images. While approaches similar to MIL with DL are not unknown in the radiological domain, such as with brain MRI for Alzheimer’s disease prediction with multiple Transformers(34), such studies do not deal with cancer prognosis and such model training approaches need to train to extract features from the raw images, whereas our study leverages FMs to sidestep that part of training, focusing only on downstream tasks alongside comparing such extractors to both handcrafted-radiomics and clinical baselines within a multicentric external validation setting.

We have shown that 2D MIL models with FM feature extraction achieve good classification performance across 2-year endpoints for HNSCC patients with performances similar to the best DL image models in previous RADCURE studies(26) for 2-year OS as well as significant stratification of patients into risk groups, indicating 2D MIL approaches with FMs as competitive for 2-year endpoint prediction as well as stratification of patients into risk groups for OS and LRC, with broader stratification for LRC than other studies employing 2D approaches for HNSCC(35). Information from multiple views of the CT did not result in consistent increases in performance or stratification, suggesting that information from coronal and sagittal views is of limited relevance when already considering the axial slices for outcome prediction. When considering the 3D FM by Pai et al.(14), performances across all 2-year endpoints were lower than in 2D models. The cause of this might be both a lower quantity of data used(36) for originally training the 3D FM when compared to other FMs like BioMedClip as well as training mostly on general abdominal and chest lesions, suggesting that FMs trained on specific sections of the body may not be optimal for prediction within other body regions. Thus, there is a need for larger CT datasets to create better-performing FMs as well as for assessment of FMs in clinically-relevant prediction tasks inside and outside of their original anatomical region to better define their applicability scope.

Handcrafted radiomics achieved generally similar or slightly lower performance for 2-year endpoint prediction and stratification compared to 2D MIL models. This performance disparity is in accordance with Gouthamchand et al.(37) where handcrafted radiomics models were shown to systematically offer slightly lower performances over DL models in HNSCC studies. The inclusion of information of not just the GTVp but also of surrounding context from the CT slice through the DL feature representations might provide information related to aspects such as tumor spread across the slice and explain the observed gains in both classification performance and stratification. While previous studies have included handcrafted radiomics information outside the GTVp(38), showcasing increased performance compared to GTVp-only handcrafted radiomics features, these studies require tuning the size of peritumoral volume to be included in contrast to DL approaches.

While statistically significant predictors in a multivariate setting, MIL scores showed positive correlation across endpoints with tumor T stage for all external cohorts; indicating that models learned patterns associated with known clinical factors. Correlations of scores to clinical parameters were shown to be endpoint-dependent as positive correlations with N-stage for 2-year FFDM were observed across all three test cohorts, which is a known risk factor for distant metastasis in HNSCC(39) while showing little to no correlations to N-stage for 2-year OS MIL scores. Increases in performance for the multimodal models when compared to the clinical baseline were observed in the subset of patients of non-HPV+ tumors, indicating potential for MIL approaches for multimodal modeling with clinical baselines. Such increases were, however, not consistent across all endpoints and test cohorts. Bias mitigation techniques for learning scores that are less redundant with known clinical factors while training for endpoint-prediction tasks(40) could be considered for future work in order to better synergize AI predictions with known clinical baselines.

The present study is not without its limitations; considered data is from publicly.available retrospective cohorts and further validation is required to extend the modeling scope beyond a proof of concept for the applicability of MIL and FM approaches in HNSCC with pretreatment CTs. Secondly, the paper does not consider handcrafted radiomics features within the peritumoral area. Thirdly, while other 3D FMs based on CT exist such as MERLIN(41) or CT-CLIP(42), they are not evaluated due to non-suitability for MIL modelling. And lastly, the study does not explore applicability of FMs as extractors in settings with low training data for endpoint prediction models.

In conclusion, the study has shown the potential of 2D MIL models with FM backbones for competitive endpoint prediction and stratification in HNSCC patients with similar or higher performance than handcrafted radiomics models as well as added performance for non-HPV+ HNSCC tumors.

## List of Abbreviations

FM: foundation model

MIL: multiple instance learning

HNSCC: head and neck squamous cell carcinoma

GTVp: primary gross tumor volume

AUROC: area under the receiver operator curve

LOR: log-odds ratio

HR: hazard ratio

OS: overall survival

LRC: locoregional control

FFDM: freedom from distant metastasis

HPV+: human papillomavirus positive

## Data Availability

Code to reproduce the results is available at https://github.com/KatherLab/HNSCC_FM_MIL_study, data used for development and validation of the paper can be accessed through request to The Cancer Imaging Archive (TCIA)

