## Supplementary Material for "End-to-end prediction of clinical outcomes in head and neck squamous cell carcinoma with foundation model-based multiple instance learning"

**Supplementary Table 1**.- Normalisation parameters for feature extraction for MIL model per extractor. Parameters are given for the three image input channels to the models. The Pai 3D FM does not have normalisation parameters beyond the windowing described in the Methods.

| **Model** | **Model type** | **Mean** | **Standard Deviation** |
| --- | --- | --- | --- |
| ResNet50 | 2D, multiview | [0.485,0.456,0.406] | [0.229,0.224,0.225] |
| SwinViT | 2D, multiview | [0.485,0.456,0.406] | [0.229,0.224,0.225] |
| BioMedClip | 2D, multiview | [0.481,0.457,0.408] | [0.269,0.261,0.276] |
| Inflated-ResNet50 | 3D | [0.485,0.456,0.406] | [0.229,0.224,0.225] |

**Supplementary Table 2**.- Radiomics feature extraction parameters for PyRadiomics extraction.

| Library | Pyradiomics 3.1.0 |
| --- | --- |
| Interpolation Method | Bilinear Spline Interpolation |
| Extraction type | 3D |
| Voxel volume objective (mm3) | 1x1x1 |
| HU intensity range | [-10,110] |
| Pad Distance | 10 |
| Intensity Discretisation Algorithm | Fixed Bin Number |
| Number of Bins | 32 |
| Weighting norm | Infinity |
| Disabled Features? | Default |
| Symmetrical GLCM | Yes |
| Alpha level GLDM | 4 |

**Supplementary Table 3.-** CT scanning parameters of the training and test cohorts. Metadata is retrieved from the downloaded DICOM files. Missingness in a variable is indicated with “Unknown”. Categorical variables are indicated with their number per category and percentage. Continuous variables are indicated by their median and IQR.

|  |  | **RADCURE train**  **(n=1464)** | **RADCURE test**  **(n=606)** | **HN1**  **(n=131)** | **HN-PET-CT**  **(n=284)** |
| --- | --- | --- | --- | --- | --- |
| **Peak tube voltage (kV)** |  | 120.0 [120.0-120.0] | 120.0 [120.0-120.0] | Unknown: 131 | 140.0 [120.0-140.0] |
| **Tube current (mA)** |  | 300.0 [300.0-490.0] | 300 [300-500.0] | Unknown: 131 | 70.0 [29.0-394.0] |
| **x-spacing (mm)** |  | 0.98 [0.83-0.98] | 0.98 [0.83-1.17] | 0.97 [0.97-0.97] | 0.98 [0.98-1.37] |
| **Convolution Kernel** | B | 3 | 24 | 0 | 33 |
|  | C | 29 | 33 | 0 | 97 |
|  | FC03 | 1 | 0 | 0 | 0 |
|  | FC04 | 949 | 519 | 0 | 0 |
|  | FC05 | 2 | 0 | 0 | 0 |
|  | FC08 | 2 | 0 | 0 | 0 |
|  | STANDARD | 478 | 30 | 0 | 61 |
|  | SOFT | 0 | 0 | 0 | 91 |
|  | Unknown | 0 | 0 | 131 | 5 |
| **Scanner type** | Aquillion | 18 | 0 | 0 | 0 |
|  | Aquilion ONE | 689 | 519 (0.0) | 0 | 0 |
|  | Brilliance Big Bore | 32 | 57 (0.0) | 0 | 0 |
|  | Discovery 610 | 0 (0.0) | 30 (0.0) | 0 | 0 |
|  | Discovery ST | 602 | 0 (0.0) | 0 | 142 |
|  | LightSpeed Plus | 123 | 0 (0.0) | 0 | 0 |
|  | SOMATOM Sensation 16 | 0 | 0 | 131 | 0 |
|  | GeminiGXL 16 | 0 | 0 | 0 | 141 |
|  | Hi-Art | 0 | 0 | 0 | 3 |
|  | Unknown | 0 | 0 | 0 | 1 |
| **Manufacturer** | GE MEDICAL SYSTEMS | 478 | 30 | 0 | 142 |
|  | Philips | 32 | 57 | 1 | 141 |
|  | TOSHIBA | 954 | 519 | 0 | 0 |
|  | SIEMENS | 0 | 0 | 131 | 0 |
|  | TomoTherapy Inc. | 0 | 0 | 0 | 3 |
| **z-spacing (mm)** | 1.5 | 0 | 0 | 1 | 43 |
|  | 2.0 | 986 | 519 | 0 | 25 |
|  | 2.5 | 478 | 30 | 0 | 0 |
|  | 3.0 | 0 | 0 | 130 | 74 |
|  | 3.75 | 0 | 0 | 0 | 143 |
|  | 5.00 | 0 | 0 | 0 | 2 |

Supplementary Methods: Model training

For each model in each fold, training was conducted for 64 epochs with a patience parameter of 15. Optimisation was performed through default STAMP parameters: ADAM optimiser with initial learning rate of 1e-3, maximum allowed learning rate of 1e-4 and momentum of 0.95. Learning rate scheduling was performed through cosine annealing. Weights and folds for CV were initialized with seed 1337. Model training and checkpointing was guided through the use of a weighted binary cross-entropy loss function with weights equal to the inverse proportion of each class. Checkpointing was based on decrease of the loss within the holdout validation set of the CV.

For every extracted CT slice or subvolume, an augmented version was also extracted consisting of a rotation (0 to 10 degrees) and a Gaussian blur applied with probability 0.5. Features of both augmented and non-augmented images were used for model training. At each epoch, either augmented or non-augmented features were passed into the model with probability 0.5. Only the features resulting from non-augmented images were then employed for performance assessment such as checkpointing during training and model deployment. Models were built using PyTorch 2.2.0. All training was performed on an NVIDIA RTX A100 48GB GPU.

**Supplementary Table 4**.- Coefficients and normalisation parameters for the baseline clinical model for 2-year OS classification. Positive class is death before 2 years. Logistic regression log-odds ratios (LOR) are given with median and 95% CI. T (T1<T2<T3<T4) and N stage are ordinal encoded (N0<N1<N2<N3).

| **Variable** | **Coefficient [95% CI]** | **p-value** | **Mean** | **St. deviation** |
| --- | --- | --- | --- | --- |
| Intercept | -1.45 [-1.71- -1.19] | **<0.001** | NA | NA |
| T  (Ref: T1) | 0.70 [0.62-0.79] | **<0.001** | NA | NA |
| N  (Ref: N0) | 0.68 [0.58-0.78] | **<0.001** | NA | NA |
| Sex  (Ref: Female) | -0.026 [-0.24- -0.19] | 0.81 | NA | NA |
| Treatment  (Ref: Radiotherapy alone) | -0.86 [-1.08- -0.64] | **<0.001** | NA | NA |
| HPV status  (Ref: Non-positive) | -1.14 [-1.35- -0.94] | **<0.001** | NA | NA |
| Age | 0.35 [0.25-0.44] | **<0.001** | 63.66 | 10.83 |

**Supplementary Table 5**.- Coefficients and normalisation parameters for the baseline clinical model for 2-year LRC classification. Positive class is recurrence before 2 years. Logistic regression log-odds ratios (LOR) are given with median and 95% CI. T (T1<T2<T3<T4) and N stage are ordinal encoded (N0<N1<N2<N3).

| **Variable** | **Coefficient [95% CI]** | **p-value** | **Mean** | **St. deviation** |
| --- | --- | --- | --- | --- |
| Intercept | -0.66 [-0.92 - -0.40] | **<0.001** | NA | NA |
| T  (Ref: T1) | 0.67 [0.57-0.77] | **<0.001** | NA | NA |
| N  (Ref: N0) | 0.26 [0.15-0.38] | **<0.001** | NA | NA |
| Sex  (Ref: Female) | -0.07 [-0.31- 0.144] | 0.55 | NA | NA |
| Treatment  (Ref: Radiotherapy alone) | -0.64 [-0.91 - -0.37] | **<0.001** | NA | NA |
| HPV status  (Ref: Non-positive) | -2.03 [-2.30- -1.76] | **<0.001** | NA | NA |
| Age | 0.048 [-0.05-0.14] | 0.32 | 62.90 | 10.71 |

**Supplementary Table 6**.- Coefficients and normalization parameters for the baseline clinical model for 2-year FFDM classification. Positive class is metastasis before 2 years. Logistic regression log-odds ratios (LOR) are given with median and 95% CI. T (T1<T2<T3<T4) and N stage are ordinal encoded (N0<N1<N2<N3).

| **Variable** | **Coefficient [95% CI]** | **p-value** | **Mean** | **St. deviation** |
| --- | --- | --- | --- | --- |
| Intercept | -2.01 [-2.31- -1.71] | **<0.001** | NA | NA |
| T  (Ref: T1) | 0.53 [0.44-0.62] | **<0.001** | NA | NA |
| N  (Ref: N0) | 0.84 [0.73-0.95] | **<0.001** | NA | NA |
| Sex  (Ref: Female) | 0.39 [0.14-0.64] | **0.002** | NA | NA |
| Treatment  (Ref: Radiotherapy alone) | -0.28 [-0.50- -0.52] | **0.016** | NA | NA |
| HPV status  (Ref: Non-positive) | -0.60 [-0.80 - -0.40] | **<0.001** | NA | NA |
| Age | 0.21 [0.11-0.31] | **<0.001** | 62.85 | 10.69 |

**Supplementary Table 7.-** Coefficients with 95% CI, p-values and normalisation parameters (mean and standard deviation) for selected radiomics features for 2-year OS. All features extracted come from the baseline CT image without filters.

| **Feature** | **Coefficient (95% CI)** | **p-value** | **Mean** | **Standard deviation** |
| --- | --- | --- | --- | --- |
| Intercept | -0.24 [-0.33- -0.15] | **<0.001** | NA | NA |
| Shape_Maximum3DDiameter | 0.69 [0.52-0.87] | **<0.001** | 54.50 | 23.24 |
| GLDM_LargeDependenceLowGreyEmphasis | 0.49 [0.23-0.75] | **<0.001** | 65.98 | 14.36 |
| Shape_VoxelVolume | 0.03 [-0.18-0.25] | 0.76 | 24783.27 | 32974.41 |
| GLSZM_LargeAreaEmphasis | -0.03 [-0.21-0.15] | 0.70 | 908099.09 | 1747762.31 |
| GLRLM_RunEntropy | -0.14 [-0.34- 0.07] | 0.18 | 13.54 | 0.33 |
| GLSZM_GreyLevelVariance | 0.12 [-0.02-0.26] | 0.095 | 3.87 | 0.41 |
| GLDM_DependenceEntropy | -0.49 [-0.81- -0.17] | **0.003** | 4.49 | 0.81 |
| GLSZM_ZoneEntropy | -0.31 [-0.46- -0.16] | **<0.001** | 4.34 | 0.43 |
| GLSZM_HighGrayLevelZoneEmphasis | -0.02 [-0.16-0.12] | 0.76 | 9.79 | 2.33 |
| GLRLM_LongRunEmphasis | 0.10 [-0.07- 0.26] | 0.26 | 13.54 | 11.84 |
| Firstorder_Minimum | -0.24 [-0.66 -0.18] | 0.27 | -9.74 | 2.21 |
| GLCM_ClusterShade | 0.05 [-0.14-0.29] | 0.097 | -0.41 | 0.67 |
| Firstorder_MeanAbsoluteDeviation | -0.13 [-0.83,0.57] | 0.36 | 15.26 | 3.64 |
| Firstorder_90Percentile | 0.22 [-0.48-0.92] | 0.54 | 74.73 | 11.90 |
| Firstorder_Kurtosis | 0.21 [0.01-0.41] | **0.041** | 4.00 | 1.13 |
| Firstorder_Maximum | -0.25 [-0.35- -0.15] | **0.005** | 109.13 | 4.23 |
| GLCM_JointAverage | 1.29 [0.44-2.15] | **0.003** | 3.15 | 0.34 |
| Firstorder_Skewness | 0.47 [0.28-0.66] | **<0.001** | -0.37 | 0.51 |
| Firstorder_10Percentile | -0.94 [-1.80- -0.85] | **0.031** | 25.33 | 12.64 |

**Supplementary Table 8.-** Coefficients with 95% CI, p-values and normalisation parameters (mean and standard deviation) for selected radiomics features for 2-year LRC. All features extracted come from the baseline CT image without filters.

| **Feature** | **Coefficient (95% CI)** | **p-value** | **Mean** | **Standard deviation** |
| --- | --- | --- | --- | --- |
| Intercept | -0.18 [-0.29 - -0.09] | **<0.001** | NA | NA |
| Shape_Maximum3DDiameter | 0.67 [0.50- 0.84] | **<0.001** | 52.08 | 21.94 |
| GLSZM_HighGreyLevelZoneEmphasis | 0.23 [0.10-0.36] | **0.001** | 9.79 | 2.34 |
| Shape_VoxelVolume | -0.17 [-0.30- -0.01] | **0.045** | 21485.99 | 29863.04 |
| GLRLM_RunEntropy | 0.42 [0.22-0.61] | **<0.001** | 3.85 | 0.42 |
| GLSZM_GreyLevelVariance | 0.31 [0.17-0.46] | **<0.001** | 2.22 | 0.33 |
| GLSZM_LargeAreaEmphasis | 0.12 [-0.01- 0.23] | 0.054 | 765770.11 | 1596580.87 |
| GLCM_ClusterShade | -0.41 [-0.56- -0.26] | **<0.001** | -0.40 | 0.66 |
| Firstorder_10Percentile | 0.71 [0.15-1.28] | **0.014** | 25.39 | 12.72 |
| GLRLM_LongRunEmphasis | 0.07 [-0.09-0.23] | 0.39 | 12.92 | 11.27 |
| GLSZM_DependenceEntropy | 0.64 [0.35-0.93] | **<0.001** | 4.54 | 0.79 |
| GLDM_ZoneEntropy | 0.30 [-0.46- -0.14] | **<0.001** | 4.32 | 0.44 |
| Firstorder_Minimum | 0.04 [-0.11-0.19] | 0.59 | -9.69 | 2.41 |
| Firstorder_MeanAbsoluteDeviation | -0.40 [-0.78- -0.01] | **0.044** | 15.29 | 3.65 |
| GLCM_JointAverage | -0.66 [-1.18 - -0.13] | **0.014** | 3.15 | 0.34 |
| Firstorder_Kurtosis | -0.33 [-0.53- -0.13] | **<0.001** | 3.99 | 1.13 |

**Supplementary Table 9.-** Coefficients with 95% CI, p-values and normalisation parameters (mean and standard deviation) for selected radiomics features for 2-year FFDM. All features extracted come from the baseline CT image without filters.

| **Feature** | **Coefficient (95% CI)** | **p-value** | **Mean** | **Standard deviation** |
| --- | --- | --- | --- | --- |
| Intercept | -0.37 [-0.47- -0.28] | **<0.001** | NA | NA |
| Shape_Maximum3DDiameter | 1.06 [0.87-1.26] | **<0.001** | 52.51 | 22.23 |
| GLDM_DependenceEntropy | -1.52 [-1.85- -1.18] | **<0.001** | 4.52 | 0.80 |
| shape_VoxelVolume | -0.59 [-0.84- -0.34] | **<0.001** | 21899.13 | 29277.66 |
| GLDM_LargeDependenceLowGrayLevelEmphasis | 0.08 [-0.17-0.33] | 0.532 | 65.48 | 14.15 |
| GLSZM_LargeAreaEmphasis | 0.15 [-0.07- 0.37] | 0.17 | 808275.35 | 1678391.13 |
| GLRLM_RunEntropy | -0.33 [-0.53- -0.13] | **0.001** | 3.85 | 0.33 |
| GLSZM_ZoneEntropy | -0.30 [-0.47- -0.13] | **0.001** | 4.32 | 0.44 |
| GLRLM_LongRunEmphasis | -0.18 [-0.34 – -0.03] | **0.024** | 13.04 | 11.08 |
| GLSZM_HighGrayLevelZoneEmphasis | -0.12 [-0.24 -0.02] | 0.083 | 9.78 | 2.36 |
| Firstorder_MeanAbsoluteDeviation | 0.90 [0.11-1.69] | **0.025** | 15.26 | 3.66 |
| Firstorder_Kurtosis | 0.37 [0.15-0.58] | **<0.001** | 4.00 | 1.13 |
| Firstorder_10Percentile | -0.13 [-0.96-0.70] | 0.77 | 25.38 | 12.69 |
| Firstorder_90Percentile | 0.11 [-0.62- 0.84] | 0.76 | 74.75 | 11.63 |
| Firstorder_Skewkness | 0.24 [0.09-0.39] | **<0.001** | -0.37 | 0.51 |

**Supplementary Table 10.-** Coefficients with 95% CI, p-values and normalisation parameters (mean and standard deviation) for selected radiomics features for 2-year OS with the non-CE training subpopulation. All features extracted come from the baseline CT image without filters.

| **Feature** | **Coefficient (95% CI)** | **p-value** | **Mean** | **Standard deviation** |
| --- | --- | --- | --- | --- |
| Intercept | -0.15 [-0.27- -0.04] | **0.011** | NA | NA |
| shape_maximum_3DDiameter | 0.63 [0.34-0.92] | **<0.001** | 54.64 | 23.17 |
| GLDM_LargeDependenceLowGrayLevelEmphasis | 0.51 [0.34-0.92] | **0.001** | 68.79 | 14.76 |
| GLDM_GrayLevelNonUniformity | 0.23 [-0.07-0.53] | 0.12 | 10577.89 | 14160.53 |
| GLSZM_GrayLevelNonUniformity | -0.10 [-0.37-0.18] | 0.45 | 71.96 | 70.85 |
| GLDM_DependenceEntropy | -0.43 [0.92-0.05] | 0.082 | 4.42 | 0.83 |
| GLRLM_RunEntropy | -0.48 [-0.83- -0.15] | **0.005** | 3.97 | 0.35 |
| GLRLM_LongRunEmphasis | 0.14 [-0.08-0.36] | 0.22 | 15.67 | 13.02 |
| Firstorder_Minimum | -0.40 [-0.74- -0.06] | **0.020** | -9.61 | 3.07 |
| GLSZM_ZoneEntropy | -0.33 [-0.55- -0.12] | **0.002** | 4.34 | 0.47 |
| Firstorder_90Percentile | 0.39 [-0.01- 0.80] | **0.057** | 72.93 | 12.15 |
| GLSZM_HighGrayLevelZoneEmphasis | -0.01 [-0.19 -0.19] | 0.93 | 9.72 | 2.69 |
| GLSZM_GrayLevelVariance | 0.16 [-0.04-0.35] | 0.11 | 2.24 | 0.47 |
| Firstorder_MeanAbsoluteDeviation | 0.01 [0.50-0.49] | 0.99 | 15.06 | 3.83 |
| Firstorder_Kurtosis | 0.02 [-0.26-0.26] | 0.99 | 4.08 | 1.21 |
| GLCM_ClusterShade | -0.06 [-0.20-0.08] | 0.39 | -0.29 | 0.67 |

**Supplementary Table 11.-** Coefficients with 95% CI, p-values and normalisation parameters (mean and standard deviation) for selected radiomics features for 2-year LRC with the non-CE training subpopulation. All features extracted come from the baseline CT image without filters.

| **Feature** | **Coefficient (95% CI)** | **p-value** | **Mean** | **Standard deviation** |
| --- | --- | --- | --- | --- |
| Intercept | -0.13 [-0.26- -0.01] | **0.038** | NA | NA |
| GLCM_ClusterShade | -0.31 [-0.43- -0.18] | **<0.001** | -0.29 | 0.65 |
| Firstorder_Maximum | -0.35 [-0.48 - -0.22] | **<0.001** | 108.87 | 5.05 |
| GLSZM_GrayLevelNonUniformity | 0.21 [-0.04-0.45] | 0.094 | 64.10 | 63.41 |
| Firstorder_Kurtosis | 0.27 [0.12-0.41] | **<0.001** | 4.04 | 1.20 |
| Firstorder_Minimum | 0.37 [0.24-0.50] | **<0.001** | -9.53 | 3.39 |
| Shape_Maximum3DDiameter | 0.15 [-0.09-0.39] | 0.21 | 52.15 | 21.65 |
| GLSZM_GrayLevelVariance | 0.57 [0.41-0.73] | **<0.001** | 2.22 | 0.47 |
| GLDM_GrayLevelNonUniformity | -0.12 [-0.32-0.07] | 0.21 | 8964.71 | 12274.20 |

**Supplementary Table 12.-** Coefficients with 95% CI, p-values and normalisation parameters (mean and standard deviation) for selected radiomics features for 2-year FFDM with the non-CE training subpopulation. All features extracted come from the baseline CT image without filters.

| **Feature** | **Coefficient (95% CI)** | **p-value** | **Mean** | **Standard deviation** |
| --- | --- | --- | --- | --- |
| Intercept | -0.32 [-0.46- -0.19] | **<0.001** | NA | NA |
| Shape_Maximum3DDiameter | 1.02 [0.73-1.32] | **<0.001** | 52.99 | 22.75 |
| GLDM_Dependence_Entropy | -0.65 [-0.91- -0.39] | **<0.001** | 4.45 | 0.83 |
| GLDM_GrayLeveNonlUniformity | -0.20 [-0.49 - -0.09] | 0.18 | 9569.25 | 13425.55 |
| GLSZM_GrayLevelNonUniformity | -0.17 [-0.45-0.10] | 0.22 | 65.93 | 65.63 |
| GLRLM_LongRunEmphasis | 0.12 [-0.09-0.33] | 0.27 | 15.04 | 11.99 |
| GLSZM_HighGrayLevelZoneEmphasis | -0.15 [-0.29- -0.01] | **0.047** | 9.73 | 2.72 |
| GLRLM_RunEntropy | -0.49 [-0.74- -0.24] | **<0.001** | 3.95 | 0.35 |

**Supplementary Table 13.-** Coefficients with 95% CI, p-values and normalisation parameters (mean and standard deviation) for selected radiomics features for 2-year OS with the CE training subpopulation. All features extracted come from the baseline CT image without filters.

| **Feature** | **Coefficient (95% CI)** | **p-value** | **Mean** | **Standard deviation** |
| --- | --- | --- | --- | --- |
| Intercept | -0.34 [-0.48- -0.18] | **<0.001** | NA | NA |
| Shape_Maximum3DDiameter | 0.72 [0.49-0.96] | **<0.001** | 54.37 | 23.30 |
| Shape_VoxelVolume | 0.06 [-0.23-0.35] | 0.41 | 24094.59 | 32913.32 |
| GLDM_LargeDependenceLowGreyLevelEmphasis | 1.36 [0.80-1.92] | **<0.001** | 63.46 | 13.51 |
| GLSZM_LargeAreaEmphasis | -0.05 [-0.31-0.21] | 0.71 | 719823.89 | 1253492.31 |
| GLSZM_GrayLevelVariance | 0.25 [0.05-0.45] | **0.013** | 2.22 | 0.35 |
| GLDM_LargeDependenceEmphasis | -0.65 [-1.40-0.11] | 0.092 | 537.45 | 75.87 |
| GLSZM_ZoneEntropy | -0.27 [-0.49- -0.06] | **0.011** | 4.35 | 0.39 |
| GLDM_DependenceEntropy | -0.25 [-0.81-0.32] | 0.391 | 4.55 | 0.77 |
| GLRLM_LongRunEmphasis | 0.11 [-0.09-0.31] | 0.270 | 11.64 | 10.28 |
| Firstorder_Minimum | -0.70 [-1.32- -0.07] | **0.029** | -9,85 | 0.88 |
| Firstorder_Maximum | -0.10 [-0.29-0.09] | 0.31 | 109.31 | 3.31 |
| GLCM_ClusterShade | 0.05 [-0.11 -0.21] | 0.56 | -0.51 | 0.65 |
| GLSZM_HighGrayLevelZoneEmphasis | -0.06 [-0.24-0.12] | 0.51 | 9.86 | 1.95 |
| Firstorder_Mean | 1.45 [0.80-2.09] | **<0.001** | 53.16 | 10.34 |

**Supplementary Table 14.-** Coefficients with 95% CI, p-values and normalisation parameters (mean and standard deviation) for selected radiomics features for 2-year LRC with the CE training subpopulation. All features extracted come from the baseline CT image without filters.

| **Feature** | **Coefficient (95% CI)** | **p-value** | **Mean** | **Standard deviation** |
| --- | --- | --- | --- | --- |
| Intercept | -0.25 [-0.38 – -0.13] | **<0.001** | NA | NA |
| Shape_Maximum3DDiameter | 0.71 [0.49-0.93] | **<0.001** | 52.03 | 22.20 |
| GLSZM_HighGrayLevelZoneEmphasis | 0.45 [0.31-0.59] | **<0.001** | 2.21 | 1.98 |
| Firstorder_Energy | -0.27 [-0.44 - -0.10] | **0.002** | 20857732100.00 | 29741940300.00 |
| GLDM_LargeDependenceEmphasis | 0.40 [0.17-0.63] | **<0.001** | 532.88 | 76.32 |
| GLSZM_GrayLevelVariance | 0.17 [0.01-0.32] | **0.046** | 4.32 | 0.35 |
| GLSZM_ZoneEntropy | -0.20 [-0.47 - -0.05] | **0.015** | 9.85 | 0.40 |
| GLRLM_LongRunEmphasis | 0.09 [-0.05-0.22] | 0.20 | 11.27 | 10.21 |

**Supplementary Table 15.-** Coefficients with 95% CI, p-values and normalisation parameters (mean and standard deviation) for selected radiomics features for 2-year FFDM with the CE training subpopulation. All features extracted come from the baseline CT image without filters.

| **Feature** | **Coefficient (95% CI)** | **p-value** | **Mean** | **Standard deviation** |
| --- | --- | --- | --- | --- |
| Intercept | -0.51 [-0.67- -0.36] | **<0.001** | NA | NA |
| Shape_Maximum3DDiameter | 0.78 [0.54-1.02] | **<0.001** | 52.09 | 21.92 |
| GLSZM_HighGrayLevelZoneEmphasis | 0.13 [-0.02- 0.27] | 0.091 | 9.82 | 1.98 |
| GLDM_LargeDependenceEmphasis | 1.61 [1.07- 2.16] | **<0.001** | 533.27 | 76.73 |
| Firstorder_Energy | -0.26 [-0.47- -0.04] | **0.019** | 20616144400.00 | 2768647700.00 |
| GLDM_LargeDependenceLowGrayLevelEmphasis | 0.29 [0.14-0.43] | **<0.001** | 63.27 | 13.55 |
| GLDM_DependenceEntropy | 0.57 [0.21-0.94] | **0.002** | 4.58 | 0.77 |
| GLSZM_ZoneEntropy | -0.45 [-0.69- -0.22] | **<0.001** | 4.32 | 3.95 |

**Supplementary Table 16**.- Performance summary for subgroup analyses and statistical comparison for handcrafted radiomics models. Models trained on a subgroup of the RADCURE training cohort (CE and Non-CE) are evaluated via the AUROC metric on the subgroups of CE or Non-CE patients on each test cohort and compared to the original model, trained on all RADCURE training patients and deployed only on those subgroups. For CE model evaluation, only the RADCURE test cohort contained CE patients. Performance differences are evaluated via a one-sided permutation test with the alternative hypothesis that performance of the model trained on the subgroups was higher than for the model trained on all the training cohort.

| **Subgroup type** | **Endpoint** | **Cohort** | **AUROC subgroup**  **[95% CI]** | **AUROC**  **all**  **[95% CI]** | **p-value** |
| --- | --- | --- | --- | --- | --- |
| CE | 2-year OS | RADCURE_test | 0.73 [0.67-0.80] | 0.73 [0.65-0.80] | 0.32 |
| CE | 2-year LRC | RADCURE_test | 0.69 [0.62-0.77] | 0.71 [0.63-0.78] | 0.73 |
| CE | 2year-FFDM | RADCURE_test | 0.70 [0.64-0.77] | 0.69 [0.62-0.76] | 0.23 |
| Non-CE | 2-year OS | RADCURE_test | 0.78 [0.62-0.90] | 0.85 [0.74-0.95] | 1.00 |
| Non-CE | 2-year OS | HN1 | 0.74 [0.63-0.85] | 0.74 [0.64-0.85] | 0.51 |
| Non-CE | 2-year OS | HN_PET_CT | 0.68 [0.57-0.79] | 0.68 [0.56-0.79] | 0.44 |
| Non-CE | 2-year LRC | RADCURE_test | 0.67 [0.45-0.88] | 0.60 [0.40-0.79] | 0.20 |
| Non-CE | 2-year LRC | HN1 | 0.66 [0.53-0.76] | 0.74 [0.61-0.84] | 0.93 |
| Non-CE | 2-year LRC | HN_PET_CT | 0.63 [0.52-0.73] | 0.64 [0.53-0.74] | 0.57 |
| Non-CE | 2-year FFDM | RADCURE_test | 0.73 [0.58-0.87] | 0.78 [0.63-0.89] | 0.86 |
| Non-CE | 2-year FFDM | HN1 | 0.66 [0.47-0.86] | 0.68 [0.52-0.83] | 0.59 |
| Non-CE | 2-year FFDM | HN_PET_CT | 0.64 [0.52-0.77] | 0.67 [0.66-0.79] | 0.95 |

**Supplementary Table 17**.- Performance summary for 2D MIL models against multiview and 3D MIL models and statistical comparison. Performance differences are evaluated via a one-sided permutation test with the alternative hypothesis that performance of the model with multiview or 3D features was higher than for the model trained with only 2D axial features.

| **Model type** | **Endpoint** | **Cohort** | **AUROC model type**  **[95% CI]** | **AUROC**  **2D MIL**  **[95% CI]** | **p-value** |
| --- | --- | --- | --- | --- | --- |
| Multiview | 2-year OS | RADCURE_test | 0.77 [0.72-0.82] | 0.75 [0.69-0.80[ | 0.15 |
| Multiview | 2-year OS | HN1 | 0.73 [0.64-0.81] | 0.77 [0.68-0.85] | 0.76 |
| Multiview | 2-year OS | HN_PET_CT | 0.77 [0.66-0.89] | 0.84 [0.77-0.90] | 0.96 |
| Multiview | 2-year LRC | RADCURE_test | 0.67 [0.60-0.74] | 0.75 [0.68-0.81] | 0.98 |
| Multiview | 2-year LRC | HN1 | 0.59 [0.46-0.71] | 0.66 [0.52-0.78] | 0.83 |
| Multiview | 2-year LRC | HN_PET_CT | 0.71 [0.59-0.79] | 0.72 [0.61-0.80] | 0.61 |
| Multiview | 2-year FFDM | RADCURE_test | 0.65 [0.58-0.72] | 0.78 [0.71-0.84] | 1.00 |
| Multiview | 2-year FFDM | HN1 | 0.68 [0.55-0.81] | 0.75 [0.55-0.92] | 0.74 |
| Multiview | 2-year FFDM | HN_PET_CT | 0.54 [0.43-0.64] | 0.71 [0.59-0.80] | 0.99 |
| 3D | 2-year OS | RADCURE_test | 0.71 [0.64-0.78] | 0.75 [0.69-0.80[ | 0.85 |
| 3D | 2-year OS | HN1 | 0.65 [0.54-0.76] | 0.77 [0.68-0.85] | 0.89 |
| 3D | 2-year OS | HN_PET_CT | 0.62 [0.51-0.74] | 0.84 [0.77-0.90] | 0.99 |
| 3D | 2-year LRC | RADCURE_test | 0.71 [0.64-0.78] | 0.75 [0.68-0.81] | 0.85 |
| 3D | 2-year LRC | HN1 | 0.63 [0.50-0.74] | 0.66 [0.52-0.78] | 0.69 |
| 3D | 2-year LRC | HN_PET_CT | 0.66 [0.56-0.75] | 0.72 [0.61-0.80] | 0.81 |
| 3D | 2-year FFDM | RADCURE_test | 0.69 [0.61-0.75] | 0.78 [0.71-0.84] | 0.99 |
| 3D | 2-year FFDM | HN1 | 0.52 [0.36-0.69] | 0.75 [0.55-0.92] | 0.92 |
| 3D | 2-year FFDM | HN_PET_CT | 0.50 [0.37-0.61] | 0.71 [0.59-0.80] | 0.98 |

**Supplementary Table 18**.- Performance summary for subgroup analyses and statistical comparison for 2D MIL models. Models trained on a subgroup of the RADCURE training cohort (CE and Non-CE) are evaluated via the AUROC metric on the subgroups of CE or Non-CE patients on each test cohort and compared to the original model, trained on all RADCURE training patients and deployed only on those subgroups. For CE model evaluation, only the RADCURE test cohort contained CE patients. Performance differences are evaluated via a one-sided permutation test with the alternative hypothesis that performance of the model trained on the subgroups was higher than for the model trained on all the training cohort.

| **Subgroup type** | **Endpoint** | **Cohort** | **AUROC subgroup**  **[95% CI]** | **AUROC**  **all**  **[95% CI]** | **p-value** |
| --- | --- | --- | --- | --- | --- |
| CE | 2-year OS | RADCURE_test | 0.75 [0.68-0.82] | 0.73 [0.67-0.79] | 0.22 |
| CE | 2-year LRC | RADCURE_test | 0.70 [0.63-0.76] | 0.74 [0.68-0.80] | 0.88 |
| CE | 2year-FFDM | RADCURE_test | 0.76 [0.68-0.82] | 0.76 [0.68-0.83] | 0.52 |
| Non-CE | 2-year OS | RADCURE_test | 0.81 [0.67-0.92] | 0.82 [0.68-0.94] | 0.67 |
| Non-CE | 2-year OS | HN1 | 0.72 [0.62-0.79] | 0.77 [0.68-0.85] | 0.85 |
| Non-CE | 2-year OS | HN_PET_CT | 0.77 [0.69-0.84[ | 0.84 [0.77-0.90] | 0.95 |
| Non-CE | 2-year LRC | RADCURE_test | 0.76 [0.57-0.92] | 0.78 [0.58-0.93] | 0.60 |
| Non-CE | 2-year LRC | HN1 | 0.56 [0.43-0.69] | 0.66 [0.52-0.80] | 0.99 |
| Non-CE | 2-year LRC | HN_PET_CT | 0.75 [0.67-0.83] | 0.72 [0.61-0.80] | 0.29 |
| Non-CE | 2-year FFDM | RADCURE_test | 0.85 [0.71-0.94] | 0.78 [0.71-0.84] | 0.61 |
| Non-CE | 2-year FFDM | HN1 | 0.73 [0.53-0.91] | 0.75 [0.55-0.92] | 0.61 |
| Non-CE | 2-year FFDM | HN_PET_CT | 0.71 [0.58-0.81] | 0.71 [0.59-0.80] | 0.48 |

**Supplementary Table 19**.- Coefficients and normalisation parameters for the clinical model for 2-year OS classification with MIL score as an extra feature for all patients. Positive class is death before 2 years. Logistic regression log-odds ratios (LOR) are given with median and 95% CI.

| **Variable** | **Coefficient [95% CI]** | **p-value** | **Mean** | **St. deviation** |
| --- | --- | --- | --- | --- |
| Intercept | -1.32 [-1.60- -1.05] | **<0.001** | NA | NA |
| T  (Ref: T1) | 0.32 [0.22-0.42] | **<0.001** | NA | NA |
| N  (Ref: N0) | 0.59 [0.49-0.70] | **<0.001** | NA | NA |
| Sex  (Ref: Female) | 0.17 [-0.06- 0.39] | 0.15 | NA | NA |
| Treatment  (Ref: Radiotherapy alone) | -0.78 [-1.01- -0.54] | **<0.001** | NA | NA |
| HPV status  (Ref: Non-positive) | -0.64 [-0.86- -0.42] | **<0.001** | NA | NA |
| Age | 0.24 [0.14-0.34] | **<0.001** | 63.66 | 10.83 |
| MIL score | 0.94 [0.82-1.05] | **<0.001** | 0.49 | 0.18 |

**Supplementary Table 20**.- Coefficients and normalisation parameters for the clinical model for 2-year LRC classification with MIL score as an extra feature for all patients. Positive class is recurrence before 2 years. Logistic regression log-odds ratios (LOR) are given with median and 95% CI.

| **Variable** | **Coefficient [95% CI]** | **p-value** | **Mean** | **St. deviation** |
| --- | --- | --- | --- | --- |
| Intercept | -0.88 [-1.09 - -0.55] | **<0.001** | NA | NA |
| T  (Ref: T1) | 0.25 [0.14-0.37] | **<0.001** | NA | NA |
| N  (Ref: N0) | 0.35 [0.23-0.47] | **<0.001** | NA | NA |
| Sex  (Ref: Female) | 0.24 [-0.01-0.49] | 0.053 | NA | NA |
| Treatment  (Ref: Radiotherapy alone) | -0.79 [-1.06 - -0.51] | **<0.001** | NA | NA |
| HPV status  (Ref: Non-positive) | -1.30 [-1.56- -1.01] | **<0.001** | NA | NA |
| Age | -0.03 [-0.14 - 0.07] | 0.51 | 62.90 | 10.72 |
| MIL score | 0.89 [0.77-1.01] | **<0.001** | 0.53 | 0.14 |

**Supplementary Table 21**.- Coefficients and normalisation parameters for the baseline clinical model for 2-year FFDM classification with MIL score as an extra feature for all patients. Positive class is metastasis before 2 years. Logistic regression log-odds ratios (LOR) are given with median and 95% CI.

| **Variable** | **Coefficient [95% CI]** | **p-value** | **Mean** | **St. deviation** |
| --- | --- | --- | --- | --- |
| Intercept | -1.38 [-1.70- -1.05] | **<0.001** | NA | NA |
| T  (Ref: T1) | 0.20 [0.07 -0.31] | **<0.001** | NA | NA |
| N  (Ref: N0) | 0.66 [0.55-0.78] | **<0.001** | NA | NA |
| Sex  (Ref: Female) | 0.36 [0.11-0.61] | **0.005** | NA | NA |
| Treatment  (Ref: Radiotherapy alone) | -0.36 [-0.59- -0.14] | **0.002** | NA | NA |
| HPV status  (Ref: Non-positive) | -0.69 [-0.90 - -0.49] | **<0.001** | NA | NA |
| Age | 0.19 [0.09-0.30] | **<0.001** | 62.85 | 10.69 |
| MIL score | 0.72 [0.56-0.87] | **<0.001** | 0.43 | 0.15 |

**Supplementary Figure 1**.- ROC curves for the three endpoints for the MIL models with 2D axial features. Random performance lines are marked with a black dashed diagonal line. The best models were built with SwinViT features for 2-year OS and 2-year LRC and the BioMedClip model for 2-year FFDM.

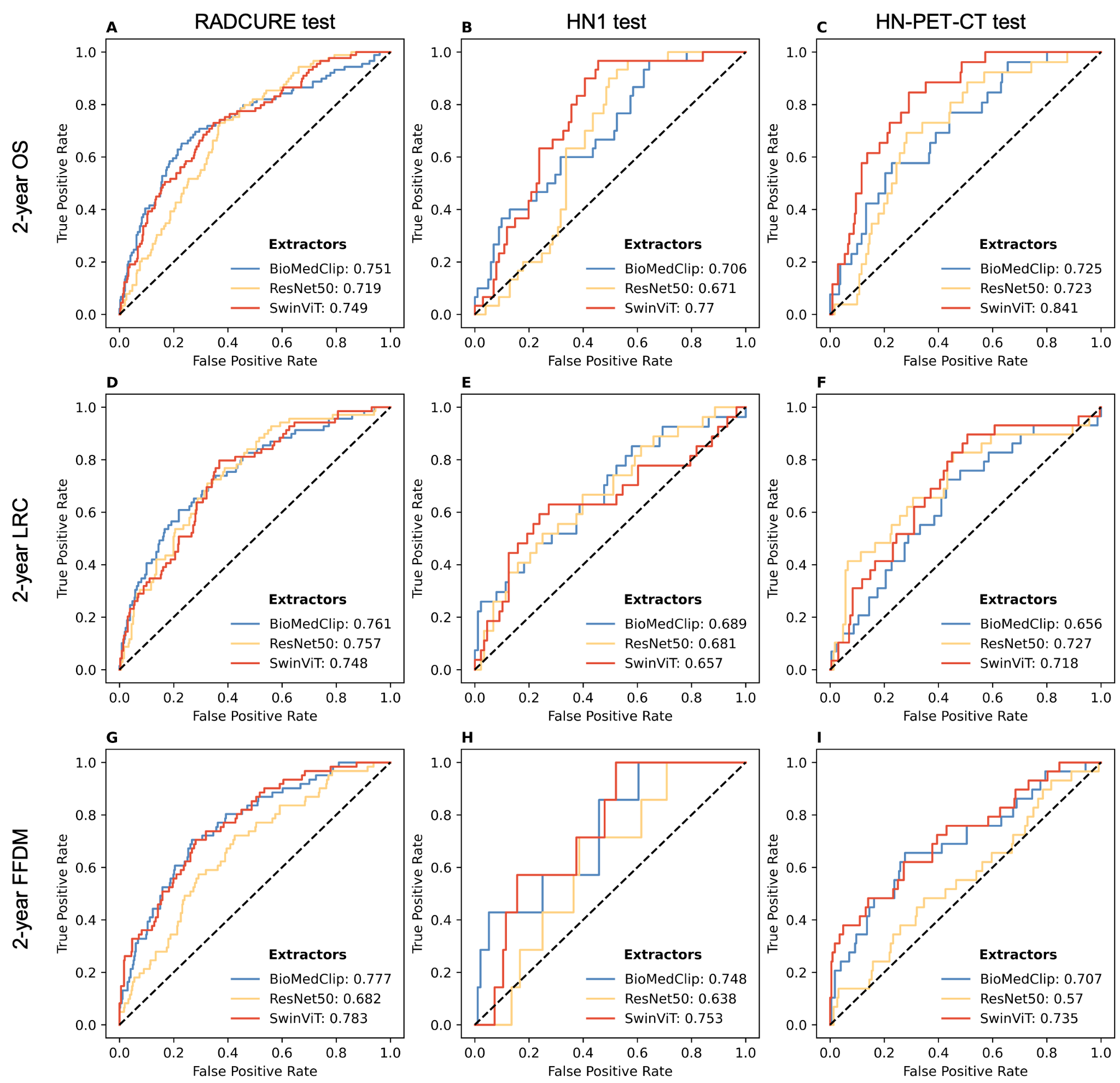

**Supplementary Figure 2**.- ROC curves for the three endpoints for the MIL models with multiview FM features. Random performance lines are marked with a black dashed diagonal line. The best models were built with SwinViT features for 2-year OS and 2-year LRC and the BioMedClip model for 2-year FFDM.

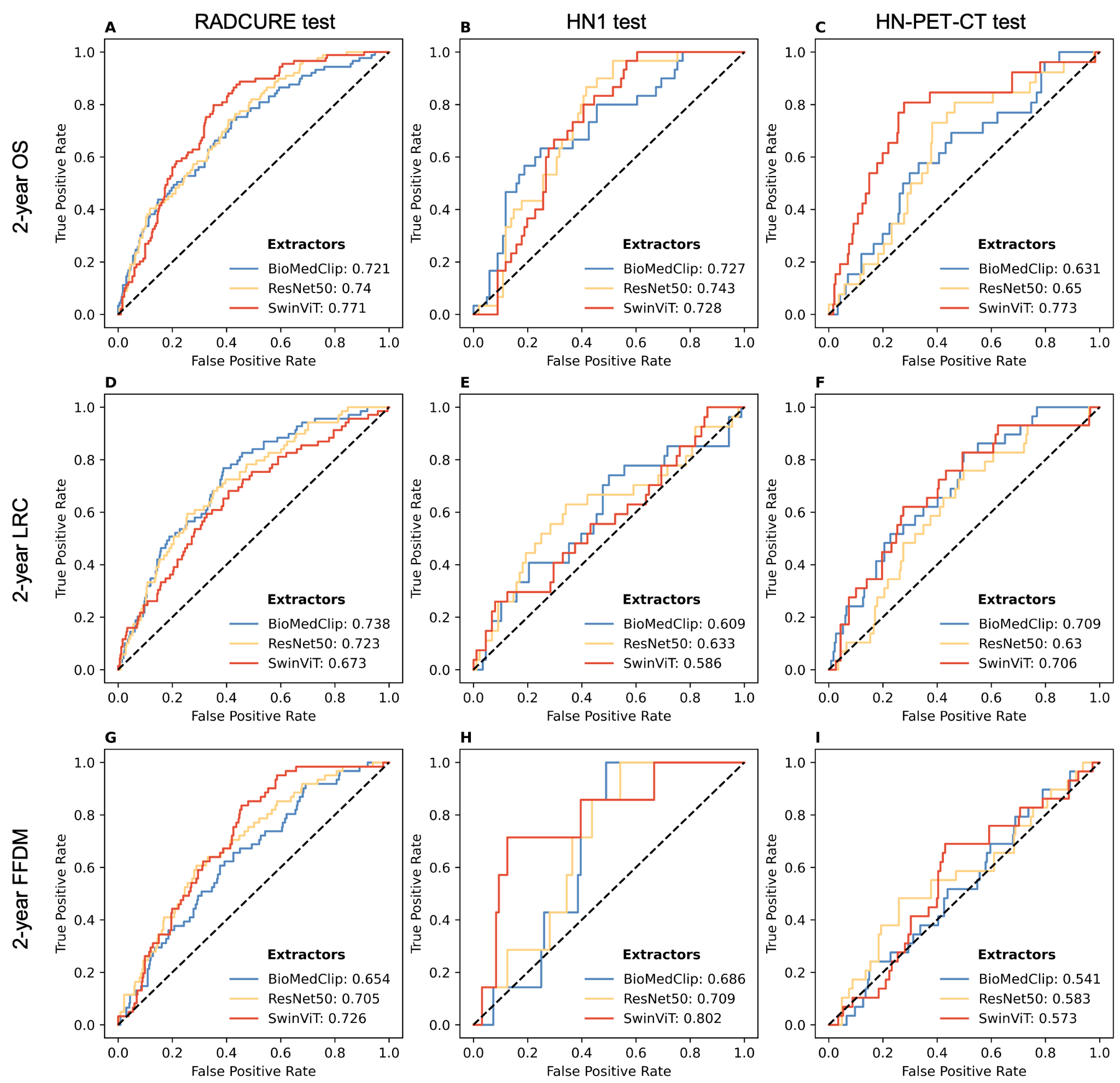

**Supplementary Figure 3**.- ROC curves for the three endpoints for the MIL models with 3D FM features. Random performance lines are marked with a black dashed diagonal line. The best models were built with Inflated-ResNet 50 for 2-year OS and 2-year FFDM and the 3D Pai FM for 2-year LRC.

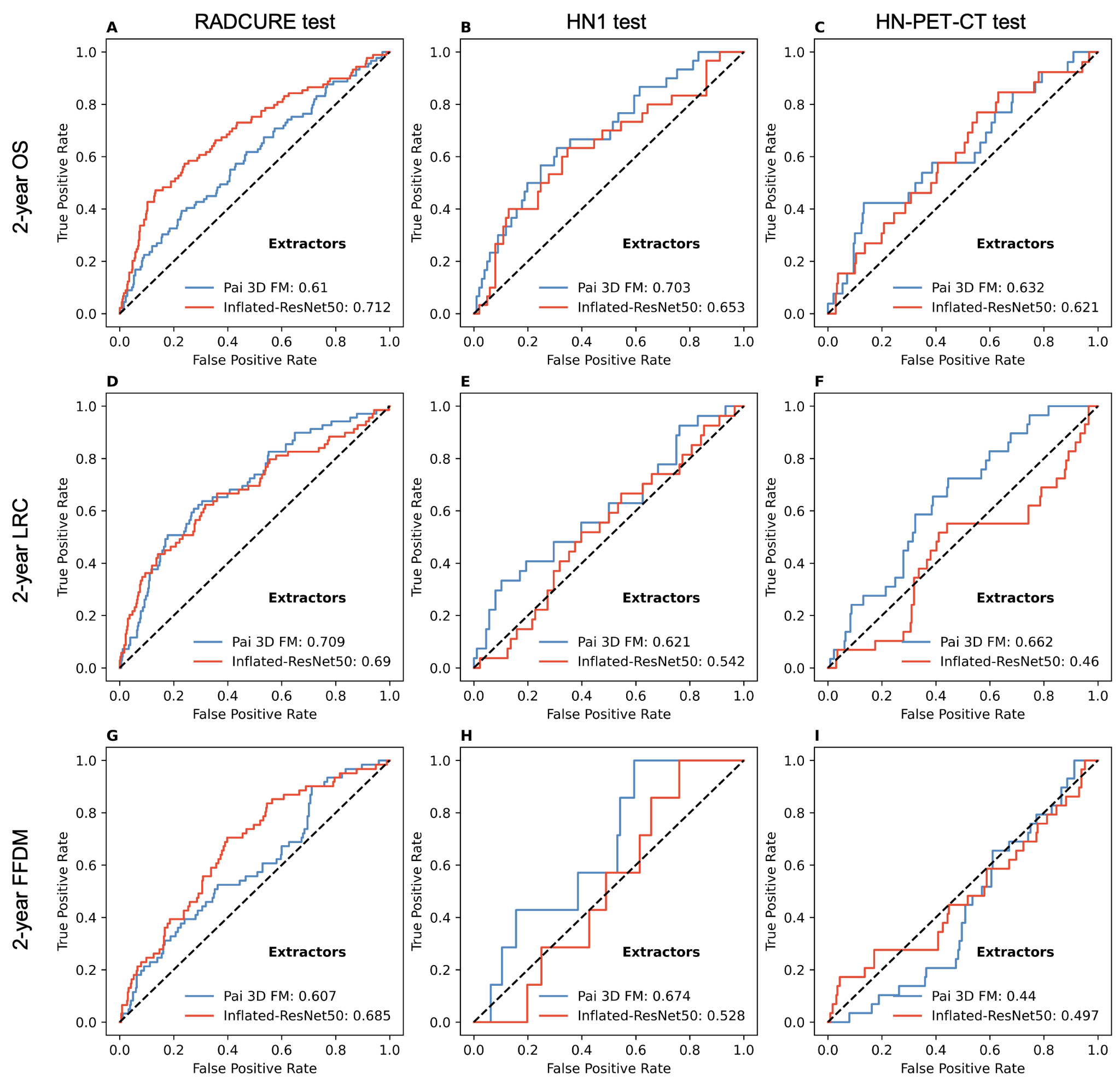

**Supplementary Figure 4.-** KM curves for stratification into high and low risk groups for the 2D MIL models for LRC (A,B,C) and FFDM (D,E,F) across the three test cohorts: RADCURE test (A,D), HN1 (B,E) and HN-PET-CT (C,F). Separation between groups is assessed through the logrank test. HR is assessed through Cox regression between the risk groups, with the low risk group used as reference.

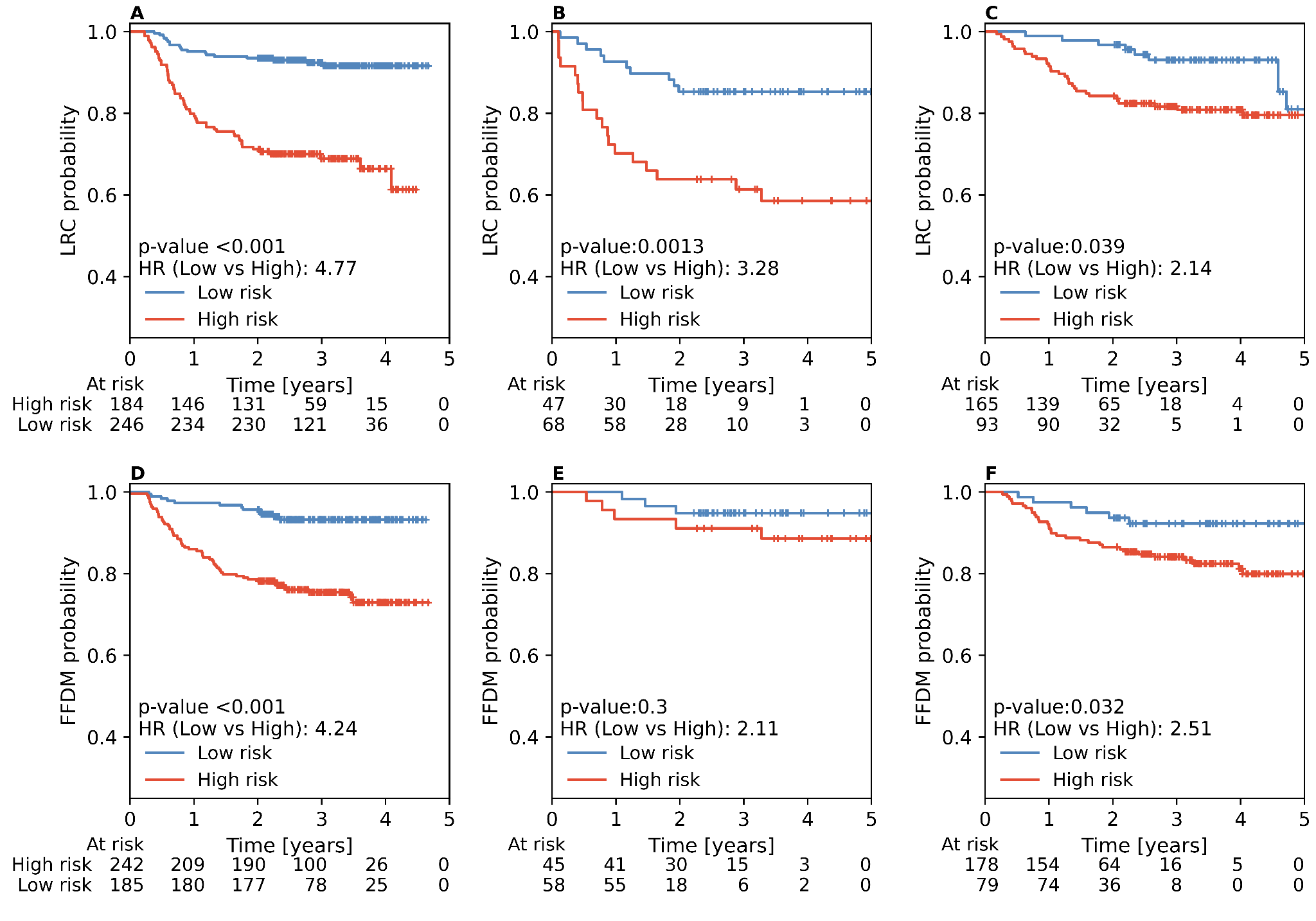

**Supplementary Figure 5.-** KM curves for stratification into high and low risk groups for the best multiview MIL models for OS (A,B,C), LRC (D,E,F) and FFDM (G,H,I) across the three test cohorts: RADCURE test (A,D,G), HN1 (B,E,H) and HN-PET-CT (C,F,I). Separation between groups is assessed through the logrank test. HR is assessed through Cox regression between the risk groups, with the low risk group used as reference. Cutoffs were 0.4660135 for OS, 0.4845263 for LRC and 0.5049221 for FFDM.

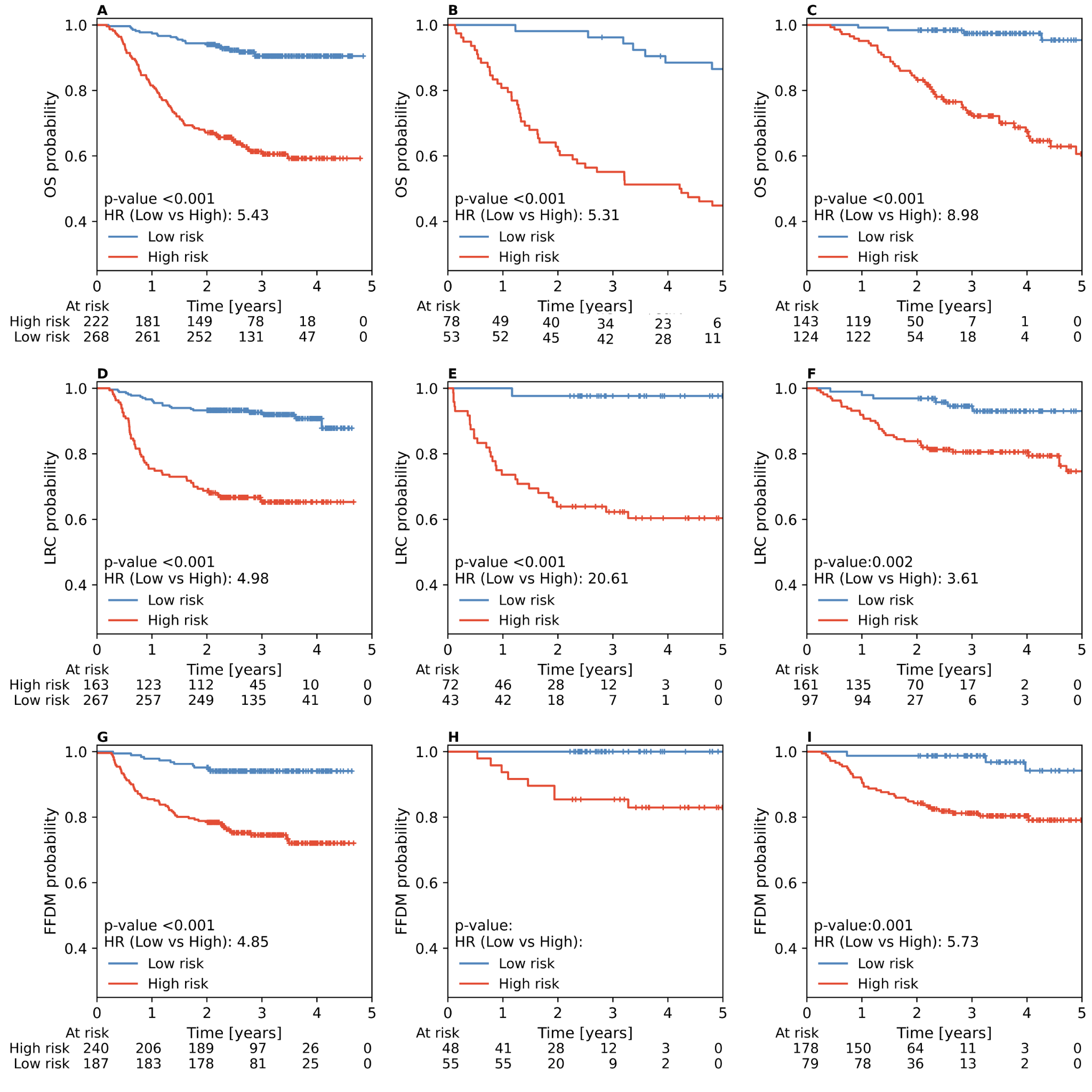

**Supplementary Figure 6.-** KM curves for stratification into high and low risk groups for the best 3D MIL models for OS (A,B,C), LRC (D,E,F) and FFDM (G,H,I) across the three test cohorts: RADCURE test (A,D,G), HN1 (B,E,H) and HN-PET-CT (C,F,I). Separation between groups is assessed through the logrank test. HR is assessed through Cox regression between the risk groups, with the low risk group used as reference. Cutoffs were 0.35400684 for OS, 0.47753704 for LRC and 0.5344289 for FFDM.

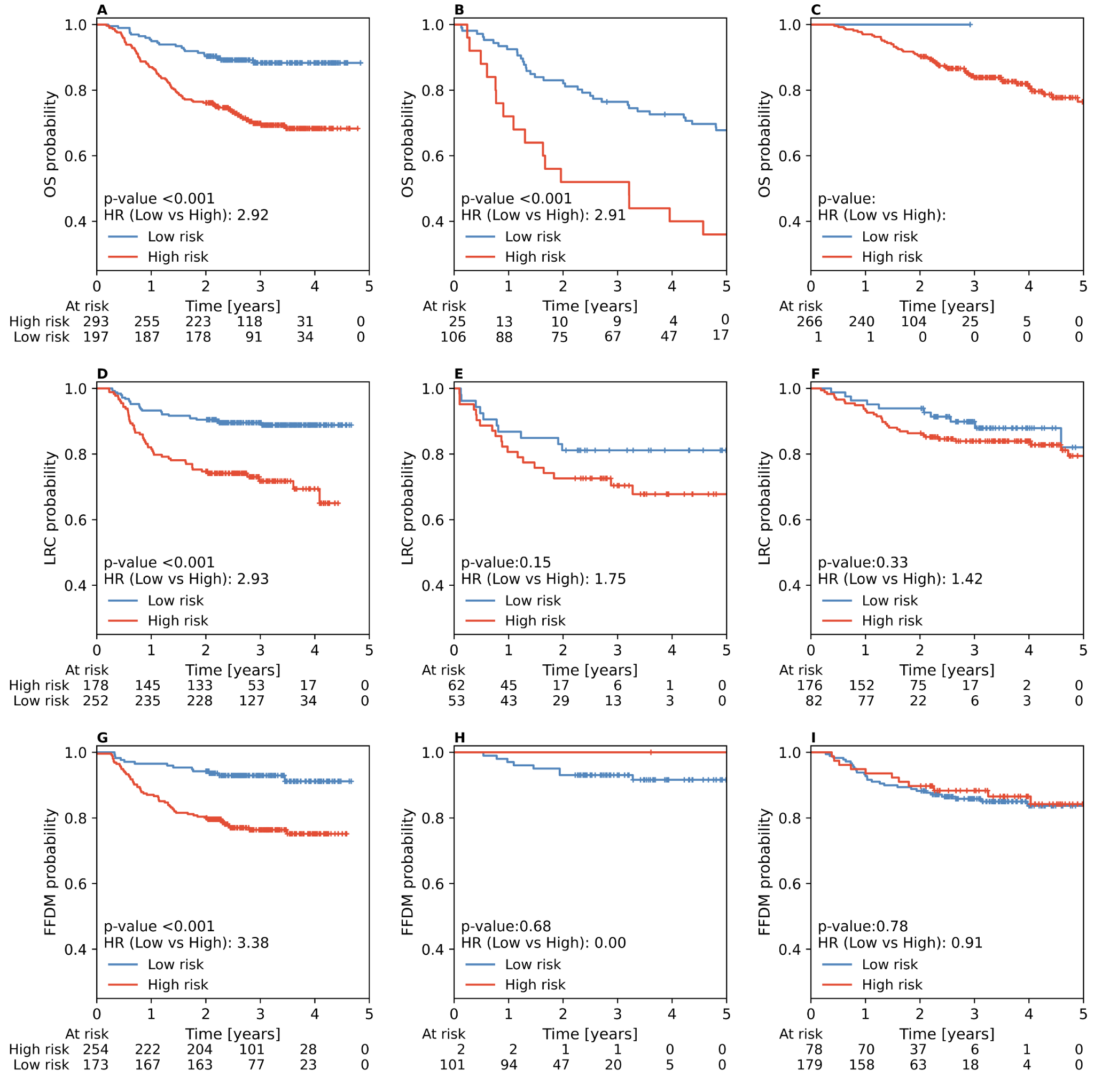

**Supplementary Figure 7.-** KM curves for stratification into high and low risk groups for the clinical baseline models for OS (A,B,C), LRC (D,E,F) and FFDM (G,H,I) across the three test cohorts: RADCURE test (A,D,G), HN1 (B,E,H) and HN-PET-CT (C,F,I). Separation between groups is assessed through the logrank test. HR is assessed through Cox regression between the risk groups, with the low risk group used as reference. Plots without HR and p-value indicate that no events were present in one of the risk groups.

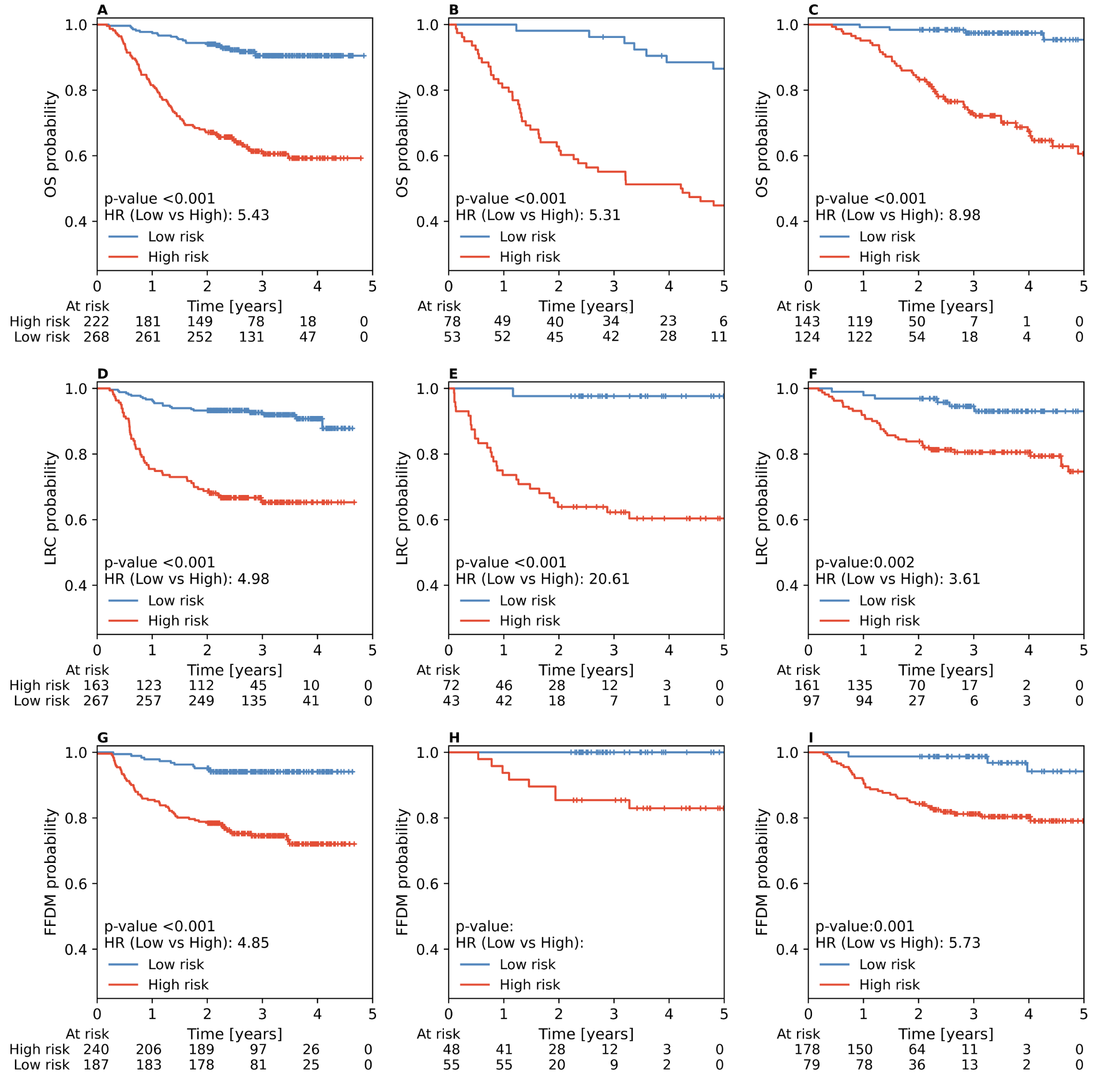

**Supplementary Figure 8.-** KM curves for stratification into high and low risk groups for the handcrafted-radiomics models for OS (A,B,C), LRC (D,E,F) and FFDM (G,H,I) across the three test cohorts: RADCURE test (A,D,G), HN1 (B,E,H) and HN-PET-CT (C,F,I). Separation between groups is assessed through the logrank test. HR is assessed through Cox regression between the risk groups, with the low risk group used as reference.

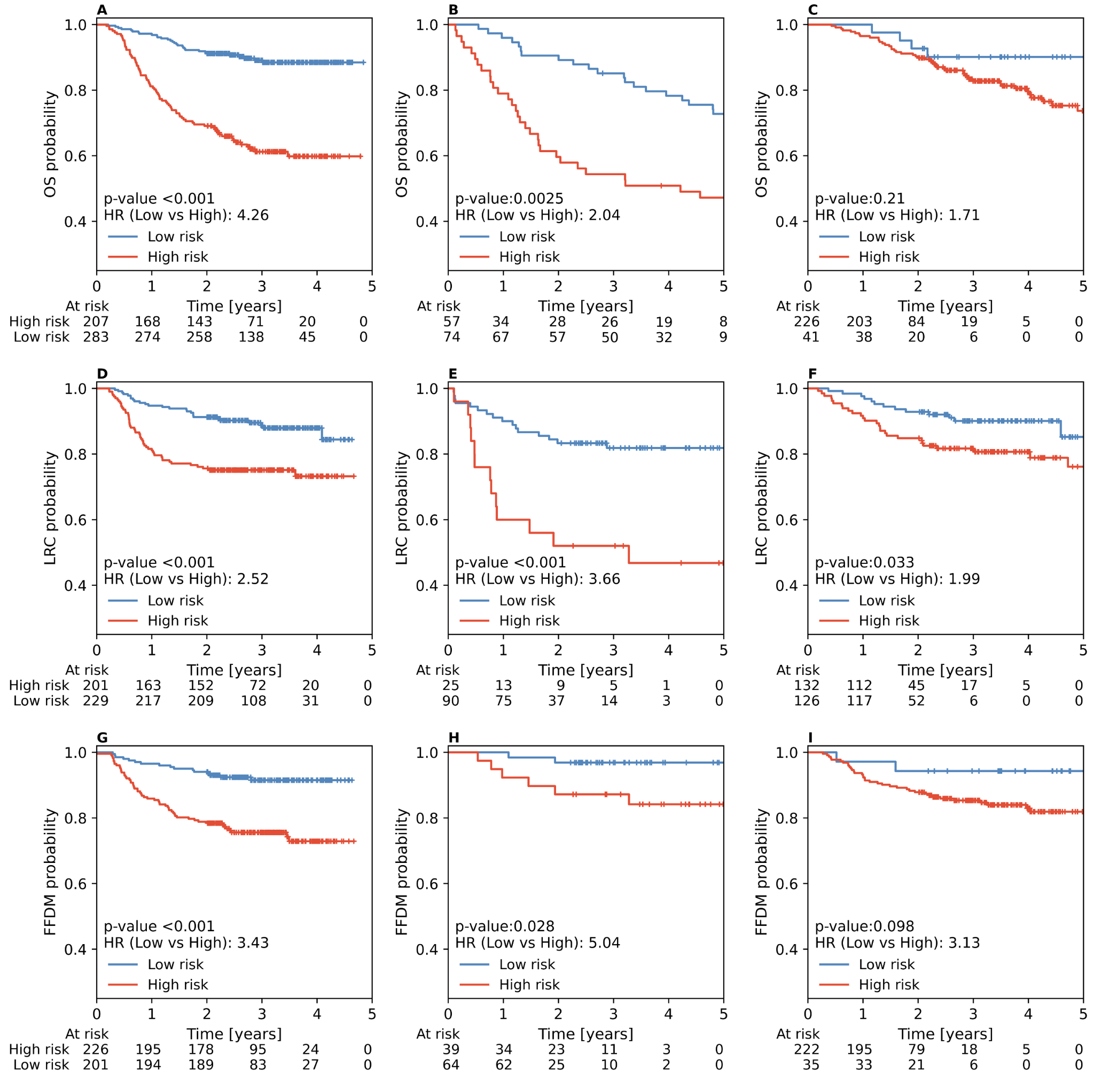
